# Comparative and Integrated Analysis of Plasma Extracellular Vesicles Isolations Methods in Healthy Volunteers and Patients Following Myocardial Infarction

**DOI:** 10.1101/2022.04.12.22273619

**Authors:** Daan Paget, Antonio Checa, Benedikt Zöhrer, Raphael Heilig, Mayooran Shanmuganathan, Raman Dhaliwal, Errin Johnson, Maléne Møller Jørgensen, Rikke Bæk, Oxford Acute Myocardial Infarction Study (OxAMI), Craig E. Wheelock, Keith M. Channon, Roman Fischer, Daniel C. Anthony, Robin P. Choudhury, Naveed Akbar

## Abstract

Plasma extracellular vesicle (EV) number and composition are altered following myocardial infarction (MI), but to properly understand the significance of these changes it is essential to appreciate how the different isolation methods affect EV characteristics, proteome and sphingolipidome. Here, we compared plasma EV isolated from platelet-poor plasma from four healthy donors and six MI patients at presentation and 1-month post-MI using ultracentrifugation, polyethylene glycol precipitation, acoustic trapping, size-exclusion chromatography (SEC) or immunoaffinity capture. The isolated EV were evaluated by Nanoparticle Tracking Analysis, Western blot, transmission electron microscopy, an EV-protein array, untargeted proteomics (LC-MS/MS) and targeted sphingolipidomics (LC-MS/MS). The application of the five different plasma EV isolation methods in patients presenting with MI showed that the choice of plasma EV isolation method influenced the ability to distinguish elevations in plasma EV concentration following MI, enrichment of EV-cargo (EV-proteins and sphingolipidomics) and associations with the size of the infarct determined by cardiac magnetic resonance imaging 6 months-post-MI. Despite the selection bias imposed by each method, a core of EV associated proteins and lipids was detectable using all approaches. However, this study highlights how each isolation method comes with its own idiosyncrasies and makes the comparison of data acquired by different techniques in clinical studies problematic.

## Introduction

Plasma extracellular vesicles (EV) are increased in number and carry altered protein, lipid and RNA cargo in the peripheral blood in many pathologies [1–6]. Analysis of plasma EV by omics approaches may provide unparalleled insight into multiple disease mechanisms and disease monitoring in patients for personalized medicine. However, it is unclear how different plasma EV isolation methods influence the plasma EV-profile or the so-called ‘EV-signature’ in patients.

Current methods for the isolation of heterogeneous plasma EV include: ultracentrifugation (UC), density ultracentrifugation, field-flow fractionation, size-exclusion chromatography (SEC), precipitation, acoustic trapping [7] and immunoaffinity capture [8]. However, unsurprisingly, laborious protocols that yield pure EV from plasma are less well favoured in large cohorts [9, 10] than protocols that are easier, lower cost, and more convenient. These methodological predilections are further impacted by the availability of stored biobank plasma, which often carry contaminating erythrocytes, immune cells, and platelets [11]. Irrespective of pre-storage processing, all plasma is a rich source of lipoproteins (apolipoprotein A and B), albumin, globulins and fibrinogens, which can co-isolate with plasma EV. The proportion of these cell-derived and non-cellular contaminants in the purified sample is method-dependent [12] and they may obscure EV associated cargo [13, 14].

Comparative isolation studies for plasma EV often assess EV size and concentration by Nanoparticle Tracking Analysis (NTA) and morphology by transmission electron microscopy (TEM). EV markers are usually evaluated using western blot or flow cytometry [15–18] . These standard analyses are often driven by the requirements of scientific bodies and consensus statements, which might be considered too prescriptive and refractory to change as our understanding of EV biology evolves [14]. However, additional assessments of how isolation methods influence the plasma EV preparation have been explored with a range of techniques, including proteomics [16, 18–21], profiling of cytokines [22], RNA integrity [23, 24], lipidomics [25], flow cytometry [15, 26] or a combination of these methods [16]. However, the wide range of plasma EV isolation and characterization techniques has evolved largely in the absence of systematic method characterization comparisons. As a consequence, the absence of an understanding of the impact of isolation techniques has led to plenty of uncertainty in relation to the interpretation of EV discoveries in clinical samples.

Here, we used platelet-free plasma from the same healthy volunteers to compare five different plasma EV isolation methods including: UC, precipitation, acoustic trapping, SEC and immunoaffinity capture using tetraspanins CD9, CD63 and CD81 (**Figure 1**). We then sought to determine how plasma EV isolation methods influence EV characteristics in a set of clinically well characterised individuals. Acute myocardial infarction (MI) is an important pathology; it is also an example of sterile inflammation where plasma EV number and composition are acutely altered [5, 6]. Plasma EV from MI patients at two different time points were isolated using the five different EV isolation methods (**Figure 1**). Integrated unsupervised analysis of all the acquired characterization data from the five different methods was used to identify the similarities and differences between each method and to highlight how each technique might influence the protein and sphingolipid composition.

**Figure 1:**
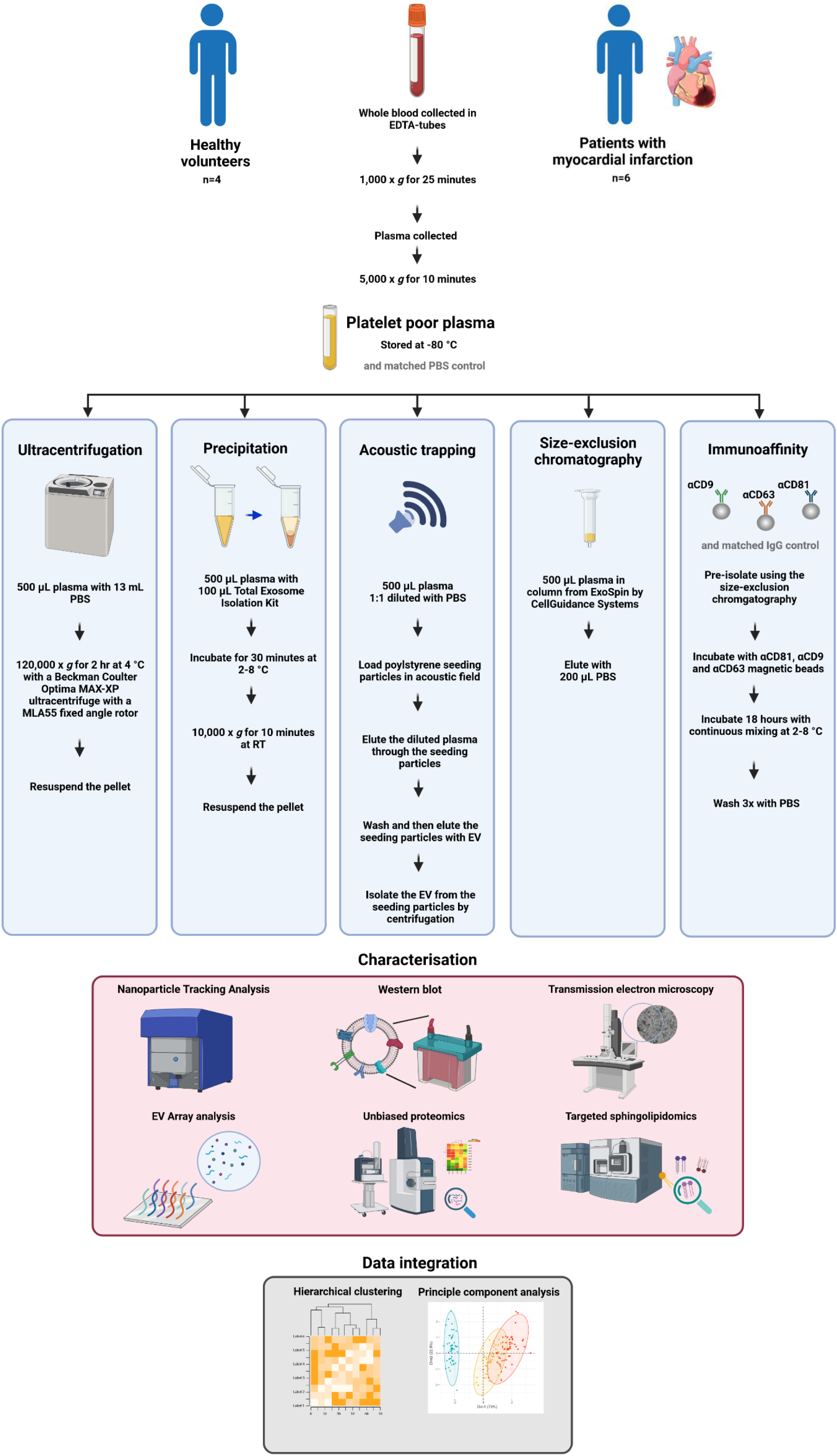
A methodological overview. Platelet poor plasma was obtained from healthy volunteers (n=4) and patients presenting with myocardial infarction (MI) (n=6) (and from the same patients 1-month post-MI) and plasma extracellular vesicles (EV) isolated using five different methods: ultracentrifugation (UC), precipitation, acoustic trapping, size exclusion chromatography (SEC) and immunoaffinity capture with a matched vehicle phosphate buffered saline (PBS) or IgG control. The plasma EV were analyzed using Nanoparticle Tracking Analysis, protein concentration, Western blot, transmission electron microscopy, a targeted EV-protein array for EV-markers CD9, CD63, CD81, ALIX, TSG101, flotillin, Annexin V and 18 other cell associated markers, untargeted proteomics (LC-MS/MS) and targeted sphingolipidomics (LC-MS/MS). The data were analyzed in insolation and following integrated hierarchical clustering and principal component analysis.

## Materials and Methods

### Healthy Volunteers and Acute Myocardial Infarction Patients

All human investigations were conducted in accordance with the Declaration of Helsinki. The Oxfordshire Research Ethics Committee (Ref: 08/H0603/41 and 11/SC/0397) approved the human clinical protocols. All healthy volunteers and myocardial Infarction (MI) patients provided informed written consent for inclusion in the study.

### Generation of Platelet Poor Plasma

Platelet-poor plasma was generated from healthy volunteers (N=4), patients presenting with MI (N=6) and from the same patients 1-month post-MI. 10-20 mL of whole blood was collected in EDTA (Greiner Bio-One, Stonehouse, United Kingdom) coated tubes and centrifuged at 1,000 x g for 25 minutes. The plasma was collected and centrifuged again for 10 minutes at 5,000 x g to produce platelet-poor plasma. The platelet-poor plasma was stored in 500 µL aliquots at -80 °C for future use.

### Isolation of plasma EV using UC

Plasma aliquots were thawed at room temperature and EV were isolated by UC by transferring 500 µL of platelet-poor plasma from healthy volunteers or 100 µL from platelet-poor plasma from MI patients to a 13.2 mL QuickSeal tube (Beckman Coulter, California, United States). The tubes were filled with a 16 G hypodermic needles (Microlance, VWR, Pennsylvania, United States) fitted to a 10 mL syringe (VWR, Pennsylvania, United States) and after plasma was injected into the tube, 13 mL of phosphate buffered saline (PBS, Thermo Fischer Scientific, Massachusetts, United States) was added. The tubes were sealed using a soldering iron (Zacro, 60 W) and centrifuged using an Optima MAX-XP ultracentrifuge (Beckman Coulter, California, United States) at 120,000 x *g* for 120 minutes at 4 °C with a MLA55 fixed-angle rotor (Beckman Coulter, California, United States) [5, 6]. The pelleted plasma EV were resuspended in 100 µL PBS or RIPA buffer (Thermo Fischer Scientific, Massachusetts, United States) for subsequent analysis.

### Isolation of plasma EV using precipitation

Plasma EV were isolated by precipitation by using the Total Exosome Isolation Kit (Invitrogen, Massachusetts, United States). 500 µL of platelet-poor plasma from healthy volunteers or 100 µL of platelet-poor plasma from MI patients was thawed at room temperature. After vortexing, 20-100 μL of the Exosome Precipitation Reagent (Total Exosome Isolation Kit (for plasma), ThermoFisher Scientific, Massachusetts, United States) was added and mixed by repeated pipetting. Samples were incubated for 30 minutes on ice, the mixture was centrifuged at 10,000 × *g* for 10 minutes by a Haraeus Fresco 17 benchtop centrifuge (ThermoFisher Scientific, ibid) at room temperature. The supernatant was removed and samples were centrifuged again at 2,000 x *g* for 2 minutes to remove residual supernatant. The pelleted plasma EV were resuspended in 100 µL PBS or RIPA buffer for downstream analysis by repeated pipetting.

### Isolation of plasma EV using acoustic trapping

Acoustic trapping of plasma for the isolation of EV was achieved by diluting plasma 1:1 with PBS as previously described [18]. Briefly, samples were loaded onto a Costar 96-well plate (Corning, New York, United States) and inserted in the AcouSort device (Version 2.0) (AcouSort AB, Lund, Sweden). Acoustic waves were produced by a waveform generator (Keysight 33210A, Keysight, California, United States) with a 9.2 V output and were directed into a borosilicate capillary acoustic trapping unit (AcouSort AB, Lund, Sweden). Following activation of the waveform generator and the acoustic trapping unit was initialised, 50 μL of 12 μm polystyrene beads (AcouSort AB, Lund, Sweden) were loaded into the acoustic trapping unit using a syringe pump (Tricontinent C2400 (Tricontinent, Fürstenfeldbruck, Germany) set to 50 μL/min. Each run consisted of 100 μL of 1:1 diluted plasma and were loaded with a syringe pump speed of 20 μL/min. After acoustic trapping, the samples were washed by aspirating 15 μL of PBS at 20 μL/min and dispensing 50 μL at 20 μL/min. After the PBS wash, the samples were eluted in 50 μL of PBS and used for downstream analysis.

### Isolation of plasma EV using size exclusion chromatography

Size exclusion chromatography (SEC) isolation of plasma EV was achieved by using the Exo-spin™ 96 (Cell Guidance Systems, Missouri, United States) [27]. Columns were equilibrated at room temperature for 15 minutes prior use. Afterwards, the columns were washed twice with 250 μL of PBS. Plasma was thawed at room temperature and 100 μL of plasma from healthy volunteers or MI patients was loaded into each column. Plasma EV were eluted by adding 200 μL of PBS to the top of the column eluted under gravity. Plasma EV were collected and stored for subsequent analysis.

### Isolation of plasma EV using immunoaffinity capture

Prior to isolation, the plasma was passed through a SEC method as described above. Exosome-Human CD9 Isolation Reagent, Exosome-Human CD63 Isolation/Detection Reagent and Exosome-Human CD81 Isolation Reagent (all Invitrogen, Massachusetts, United States) were used to capture plasma EV. The mixture was created by resuspending each bead solution and mixing by repeated pipetting. Control IgG-isotype (10400C, Thermo Fischer Scientific, Massachusetts, United States) matched beads were conjugated according to the instructions of the Dynabeads Antibody Coupling Kit (Invitrogen, Massachusetts, United States). After SEC pre-isolation, the samples were incubated with 80 μL of the combined CD9, CD63 and CD81 beads (end concentration 1 x 10^7^ / mL, equal quantity of each bead mixture) or equally concentrated IgG control beads at 4 °C for 18 hours under continual rotation by a vertical rotor (Grant Bio, Essex, United Kingdom). After incubation, the beads were pelleted for 5 minutes at room temperature using a Dynal Magnet (Invitrogen, Massachusetts, United States) and the supernatant was collected for subsequent analysis. Following pelleting, the samples were washed three times with 200 μL of PBS and magnetic beads were pelleted as described above. Following the washes, the beads were resuspended in RIPA buffer and supernatant was separated by using a 2,000 x *g* spin and collected for subsequent analysis.

### Nanoparticle Tracking Analysis

Plasma EV size distribution and concentration were determined by Nanoparticle Tracking Analysis (NTA) using a Zetaview device (Particle Metrix, Inning am Ammersee, Germany) as previously described [5, 6]. Prior to injection into the sample chamber, samples were diluted in PBS. The Zetaview measured the sample chamber from 11 positions in 2 cycles. The settings were set at sensitivity 80, fame 30 and shutter speed 100. Silica 100-nm microspheres (Polysciences Inc., Philadelphia, United States) were used to quality check the instrument performance routinely. The particle concentration per method was calculated as the change (Δ) compared to the the control sample, in which PBS replaced the plasma sample.

### Protein concentration

Protein concentration was determined by bicinchoninic assay (BCA) (Thermo Fischer Scientific, Massachusetts, United States). A standard curve with Bovine Serum Albumin (Thermo Fischer Scientific, Massachusetts, United States) was used to calculate the protein concentration. Isolated EV were diluted 1:2 or 1:6 with RIPA buffer and needle sonicated by a SonoPuls HD2070 (Bandelin, Berlin, Germany) at 40% power for 10 seconds. 25 μL of sonicated sample or standard was incubated in duplicate with 175 μL of a 25:1 ratio between Reagent A and Reagent B (Thermo Fischer Scientific, Massachusetts, United States) and incubated at 37 °C for 30 minutes. After incubation, absorbance was measured at 562 nm using a plate reader (FLUOstar Omega plate reader, BMG Labtech, Aylesbury, United Kingdom).

### Transmission Electron Microscopy

Transmission electron microscopy (TEM) of the isolated EV was conducted as previously described [6]. Briefly, grids (300 mesh Cu carbon film) were glow discharged for 20 seconds at 15 mA (Leica EM ACE 200). The isolated plasma EV samples were added to the grid for 2 minutes, blotted, stained with 2% uranyl acetate for 20 seconds, blotted and allowed to air dry. Images were acquired on a 120 kV Tecnai 12 TEM (Thermo Fischer Scientific, Massachusetts, United States) equipped with a OneView digital camera (Gatan, California, United States). TEM images of control samples, in which the isolation method was run with PBS in the place of the plasma, were obtained for each method. Immunoaffinity-based isolated EV were fixed with 1.6% glutaraldehyde in PBS for 1 hour at 4 °C. Following fixation, the beads were fixed in 4% agarose with PBS. The agarose was cut into small cubes (1-2 mm3). Sections were collected onto 200 mesh Cu grids and imaged using a Gatan OneView camera with a FEI Tecnai 12 TEM at 120kV.

### Proteomics

Isolated plasma EV were processed for proteomics as previously described [28, 29]. In short, the samples were reduced in 5 mM dithiothreitol for 30 minutes at room temperature. Subsequently, the samples were alkylated with 20 mM iodacetamide for 30 minutes and precipitation using chloroform-methanol precipitation. Quantified protein groups with □≥2 unique peptides were included in the comparison of proteomes. The LFQ values for each protein group was deducted by the LFQ from control samples per isolation method. Protein abundance was normalised by Log-transformation. Gene Ontology (GO) analysis of the protein groups [30] was conducted by using Genontology.org [30] and the data were extracted on 19-07-2021. P-values were corrected for false discovery rate (FDR) and significance was set at p<0.05. Fisher exact tests were conducted using R on published databases EVpedia [31], Vesiclepedia [32] and Exocarta [33].

### EV-Array

Isolated plasma EV were analysed by a targeted EV-Array, which has been described previously [6, 34]. In short, a protein microarray plate was generated with the following antibodies: CD146 (P1H12), Flotillin-1, TSG101 (Abnova, Taiwan), CD9, CD81 (Ancell corporation, Minnesota, United States); CD16 (3G8, BD Biosciences, California, United States); Alix (3A9), VEGFR2 (7D4-6; Biolegend, California, United States); CD63 (Bio-Rad, California, United States); ICAM-1 (R6.5, eBioscience, California, United States); Endoglin (LSbio, Washington, United States); Tissue factor (323,514), VCAM-1 (HAE-2Z), Thrombomodulin (501733), CD31 (AF806, R&D Systems, Minnesota, United States), VE-Cadherin (AF938, R&D Systems, Minnesota, United States). After blocking with the blocking buffer (50 mM ethanolamine, 100 mM Tris, 0.1% SDS, pH 9.0) for 30 minutes, the wells were emptied, and the plate was dried for 5 hours and sealed. The samples were incubated in the antibody coated microarray plate overnight at 2-8 °C. Following a wash, each well was incubated with a 100 µL of a detection antibody cocktail (biotinylated anti-human-CD9, - CD63 and -CD81 (Ancell, Minnesota, United States). After another wash, 100 µL streptavidin-Cy3 Life Technologies, Massachusetts, United States) diluted 1:3,000 was added to each well and incubated for 30 minutes. The plate was scanned using a sciREADER FL2 microarray scanner (Scienion AG, Berlin, Germany), at 535 nm and an exposure time of 2,000 milliseconds. For each protein the control, PBS sample value was subtracted from the result.

### Western blot

Isolated plasma EV or controls were lysed using RIPA buffer with protease and phosphatase inhibitors PhosSTOP (Roche, Basel, Switzerland) and cOmplete (Roche, Basel, Switzerland) and were needle sonicated by a SonoPuls HD2070 (Bandelin, Berlin, Germany) at 40% power for 10 seconds as previously described [5, 6]. Following sonication, samples were incubated at 95 °C for 5 minutes to reduce. 8 µg of protein was combined with NuPage LDS sample buffer (4x) agent (Invitrogen, Massachusetts, United States). The samples were loaded onto a 4-12% bis-tris gradient gel (NuPAGE 4-12% Bis-Tris Protein Gel; 1.5 mm (ThermoFisher Scientific, Massachusetts, United States) with Amersham ECL Full Range ladder (Cytiva Life Sciences, Massachusetts, United States). Separated samples were transferred to a nitrocellulose membrane (Amersham Proton 0.2 μm, GE Healthcare, Illinois, United States) and blocked for non-specific binding in 5 % skimmed milk powder (Marvel Original, New York, United States) in 0.5% PBS-tween 20 (Sigma-Aldrich, Missouri, United States) for 1 hour. Membranes were incubated with primary antibodies overnight: ALIX (ab117600, Abcam, Cambridge, United Kingdom) (1/1,000 dilution), CD63 (EXOAB-KIT-1, System Biosciences, California, United States) (1/1,000 dilution), ApoB (Ab139401, Abcam, Cambridge, United Kingdom) (1/20,000 dilution), albumin (MAB1455, R&D Systems, Minneapolis, Canada) (1/8,000 dilution), ApoA-I (Mab36641, R&D systems, Minneapolis, Canada) (1/8,000 dilution) and H3 (D1H2, Cell Signalling Technology, Massachusetts, United States) (1/1,000 dilution) 5 % milk in PBS-tween (PBS-T). Membranes were washed three times with PBS-T and incubated with secondary-horse radish peroxidase (HRP) conjugated antibodies (1/20,000 α-mouse W402B or 1/50,000 α-rabbit W401B, Promega, Wisconsin, United States) for 1 hour. The membranes were washed once again with PBS-T before incubating them with enhance chemiluminescence substrate (Pierce ECL, ThermoFisher Scientific, Massachusetts, United States) for imaging (Bio-Rad ChemiDoc MP Imaging system, California, United States).

### Sphingolipidomics

Sphingolipids were determined as previously described [35]. 25 μL of each sample were combined with 10 μL solution containing labelled sphingolipid internal standards in LC-MS Methanol (Honeywell, North Carolina, United States) was added to each sample. This was followed by the addition of 100 μL of methanol to each sample. Samples were then vortexed for 30 seconds and sonicated for 15 minutes in a Fisherbrand Ultrasound bath S60 (Fischer-Scientific, Massachusetts, United States) with ice. Next, samples were centrifuged at 12,000 x *g* during 15 minutes at 6LC. Finally, 80 μL of the supernatant were transferred to an LC-MS amber vial (Waters, Wilmslow, United Kingdom) equipped with a 150 μL insert. Samples were randomized by time point of the individual within an extraction method. Samples were analyzed on an Acquity UPLC coupled to a Xevo TQ-S Mass Spectrometer (Waters, Wilmslow, United Kingdom) as previously described.

### Bioinformatics and Integrated analysis

Integrated analysis was conducted by combining data from the NTA, protein concentration, EV-Array, proteomic and sphingolipidomic analysis. The data was normalized per row and log transformed. Following data normalization, a principal component analysis was conducted in R 4.0.0 [37] using the *factoextra* [38] and *FactoMineR* [39] packages. For visualization *ggplot2* [40] was used. Each of the principal components were then used to create a heatmap by extracting the values from the eigenvalues for each of the principal components and processing them in *pheatmap* [41]. The data was clustered by using double hierarchical clustering using the *pheatmap* R package.

### Data availability

All data produced in the present study are available upon reasonable request to the Corresponding Author.

### Statistical analysis

Data was plotted as mean with standard deviation. Normality of the data was confirmed using QQ plot and D’Agostino-Pearson normality test. For paired analysis, at the two time points, or for two independent groups, paired or unpaired Students T-test were used respectively (GraphPad Prism 9). Correlation analysis was carried out using linear Pearson regression analysis (Graphpad Prism 9). One-way ANOVA with Bonferroni correction post-hoc tests were used for analyses with >3 independent groups. The proteomic and lipidomic data was analyzed using a Kruskal-Wallis statistical test with Bonferroni post-hoc tests. p values <0.05 were considered significant.

## Results

### Plasma EV number and size is influenced by the isolation method

The concentration of isolated EV (expressed as delta over a matched PBS or and IgG control sample) determined by NTA differed per method (UC 9.5 x 10^9^ ± 1.8 x 10^9^ EV / mL, precipitation 6.1 x 10^11^ ± 2.7 x 10^11^ EV / mL, acoustic trapping 6.4 x 10^9^ ± 2.8 x 10^9^ EV / mL, SEC 5.5 x 10^9^ ± 1.9 x 10^9^ EV / mL and immunoaffinity 2.8 x 10^10^ ± 7.1 x 10^9^ EV / mL, **Figure 2A**). In agreement with previously published studies, precipitation yielded significantly higher numbers of particles / mL compared to UC [15, 36] and SEC [15, 37] (both p<0.01). Plasma EV number, isolated by precipitation, were also higher than the concentration acquired by acoustic trapping or by immunoaffinity capture (both p<0.01) (**Figure 2A**). The size and concentration distribution for each biological replicate exhibited uniformity within each isolation method and the different isolation methods did give a similar size distribution profile overall, which ranged from 15 nm (**Figure 2B**). However, the mean size of EV isolated by UC was significantly higher compared to the other methods (UC 143.7 ± 3.4 nm, versus precipitation 94.2 ± 3.9 nm, acoustic trapping 81.2 ± 3.9 and SEC 87.2 ± 1.5, p<0.01 all) (**Table 1**).

**Figure 2:**
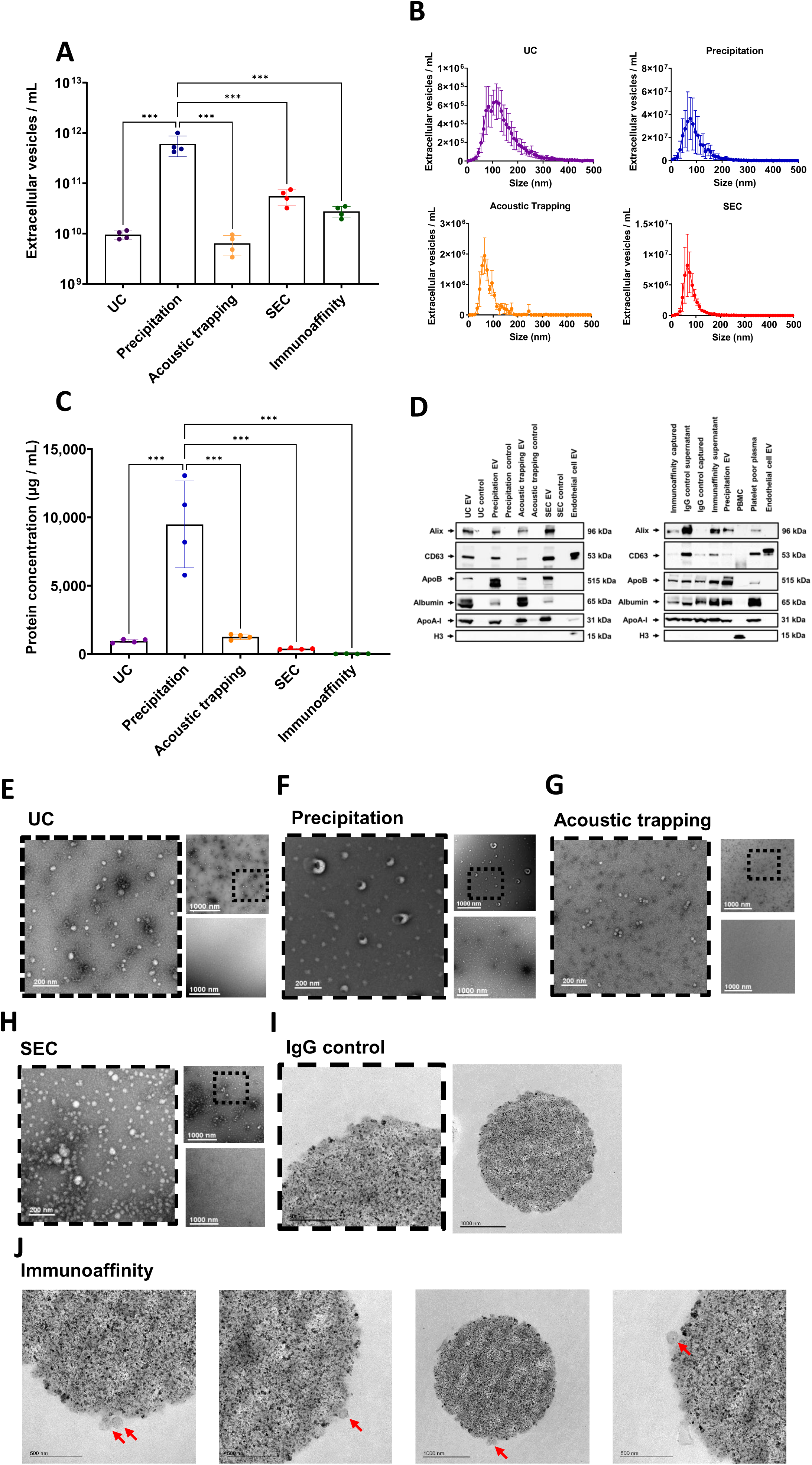
Plasma EV characterization using different isolation methods. **(A)** Total plasma extracellular vesicles (EV) / mL concentrations and **(B)** size and concentration distribution profiles were obtained by Nanoparticle Tracking Analysis (NTA) using ultracentrifugation (UC), precipitation, acoustic trapping, size exclusion chromatography (SEC) and immunoaffinity capture (n=4). Values are presented as a delta compared to a vehicle control or IgG control. Scale is logarithmic. **(C)** Protein concentration of plasma EV using UC, precipitation, acoustic trapping, SEC and immunoaffinity capture (n=4). Values are presented as a delta compared to a vehicle control or IgG control. **(D)** Western blot of plasma EV derived from UC, precipitation, acoustic trapping, SEC and immunoaffinity capture versus controls using EV markers ALIX and CD63, lipoprotein contaminants apolipoprotein B (ApoB), and apolipoprotein A-I (ApoA-I), plasma contaminant albumin and cellular contaminant histone H3. Endothelial cell EV and Peripheral blood mononuclear cells (PBMCs) were used for H3 positive controls. **(E-J).** Transmission electron microscopy (TEM) images of isolated plasma EV from UC, precipitation, acoustic trapping, SEC and immunoaffinity capture versus controls. Each sub panel contains a zoomed-in image (left image), an overview image (top right) and a control vehicle image (bottom right). For the immunoaffinity bead capture images, the red arrows indicate EV particles. The scale bar is 200 nm for the zoomed images surrounded by a dashed line and 1000 nm for the overview images and **J**. Values in **A** and **C** are group average ± standard deviation (SD). Data are group average ± standard deviation (SD) (n=6). Data was analyzed by one-way ANOVA with post-hoc Bonferroni correction. ***p<0.001.

**Table 1:**
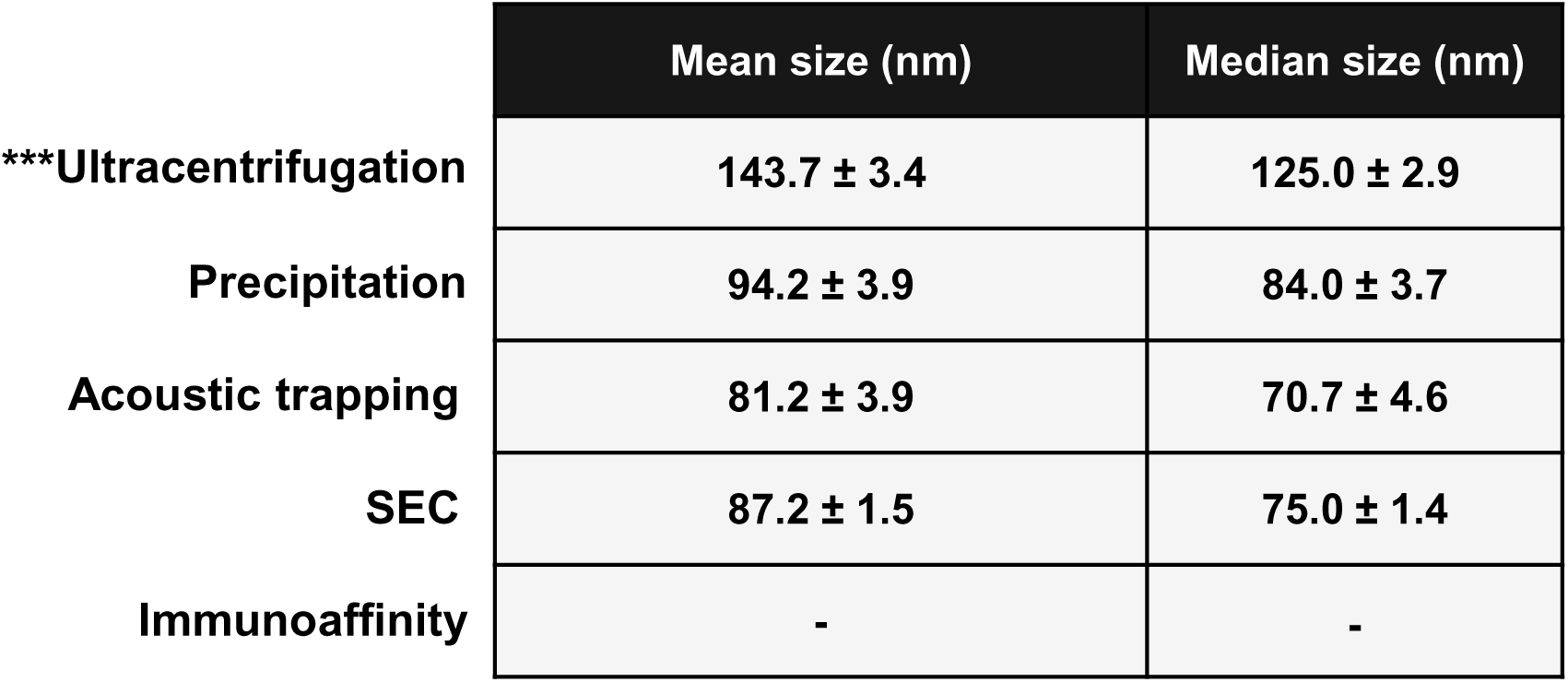
Nanoparticle Tracking Analysis showing the mean and median size of plasma-EV. isolated by ultracentrifugation (UC), precipitation, acoustic trapping, size-exclusion chromatography (SEC). Immunoaffinity capture can not be acquired. Data are group averages ± standard deviation (SD) and were analyzed by One-way ANOVA with post-hoc Bonferroni correction. ***p<0.001. (n=4).

### Plasma EV protein concentration

The protein concentration (expressed as the change over a matched PBS or and IgG control) also differed across the EV isolation methods: UC 982 ± 113 µg / mL, precipitation 9,478 ± 3,174 µg / mL, acoustic trapping 1,211 ± 141 µg / mL, SEC 382 ± 51 µg / mL and immunoaffinity 33 ± 12 µg / mL (**Figure 2C**). EV generated by precipitation had a significantly higher protein concentration compared to UC, acoustic trapping, SEC or immunoaffinity (all p<0.01, **Figure 2C**). The purity of EV isolation can be estimated by calculating a ratio of EV number (EV / mL) to protein concentration (µg / mL) [44]. The EV purity ratio differed between isolation method: UC 9.0 x 10^6^ ± 1.4 x 10^6^; precipitation 4.7 x 10^8^ ± 1.5 x 10^8^; acoustic trapping 2.1 x 10^7^ ± 7.6 x 10^6^; SEC 6.8 x 10^6^ ± 4.0 x 10^6^ and immunoaffinity capture 3.1 x 10^8^ ± 1.0 x 10^8^. Both the immunoaffinity and precipitation methods achieved a significantly higher EV purity ratio when compared to UC, SEC or acoustic trapping (p<0.01 compared to all methods) (**Supplemental Figure 1**).

### Plasma EV protein characterisation by Western blot

As NTA is unable to distinguish between plasma EV and similarly sized protein aggregates and lipoproteins, we analysed the isolated plasma EV obtained from each method for a number of markers by western blotting. ALIX and CD63 were included as the EV markers. For the plasma lipoprotein contaminants, we measured ApoB and ApoA-I. Histone H3 for cellular contaminant by western blot and albumin was also included in the set. Each isolation method showed the presence of EV markers ALIX and CD63 (**Figure 2D**). Apolipoproteins (ApoA-I and ApoB) were also present in all isolation methods. All methods were negative for markers of cellular contamination by histone H3. Surprisingly, the IgG isotype control for immunoaffinity capture using CD9, CD63 and CD81 also showed the presence of EV markers ALIX, CD63 and ApoB, albumin, ApoA-I but histone H3 was absent (**Figure 2D**).

### Plasma EV morphology by TEM

To determine the morphology of isolated plasma EV, we undertook TEM for the five different plasma EV isolation methods versus a matched PBS control or an IgG control for immunoaffinity capture beads. TEM analysis showed EV-like particles for each isolation method and an absence of EV-like particles in their respective PBS isolation controls (**Figure 2E-H**). Immunoaffinity beads CD9, CD63 and CD81 were embedded in agar and sectioned to visualise the bead surface for EV-like structures versus the IgG control. IgG control showed no EV-like particles present (**Figure 2I**). Immunoaffinity capture using CD9, CD63 and CD81 beads showed intact EV-like particles captured on the bead surface (**Figure 2J**).

### Plasma EV compositional analysis using a high throughput protein EV-Array

The results detailed above provide evidence for the relative success of each method to yield plasma EV and assessment of relative plasma EV purity [39]. However, these techniques do not readily distinguish the abundance of specific plasma EV populations carrying, for instance, specific cell-associated markers, or easily allow quantitative assessment of the abundance of contaminating lipoproteins. Thus, we probed the composition of the plasma EV isolated from each method for general EV markers CD9, CD63, CD81, ALIX, TSG101, Flotillin 1, Annexin V and a panel of 18 cell associated markers, which may distinguishes EV from platelets, endothelial cells, immune cells, muscle and lipoprotein contaminants apolipoprotein E (ApoE) and apolipoprotein H (ApoH) using a validated high throughput EV-protein antibody array (**Figure 3**) [34]. We utilised a matched PBS control for UC, precipitation, acoustic trapping and SEC and an IgG control for immunoaffinity capture. For the EV-associated proteins, CD9 and CD81, they were significantly higher in plasma EV isolated by UC compared to the other methods (p<0.01 for both). Annexin V was significantly higher in the acoustic trapping samples compared to SEC and immunoaffinity capture (p<0.01 and p<0.05, respectively).

**Figure 3:**
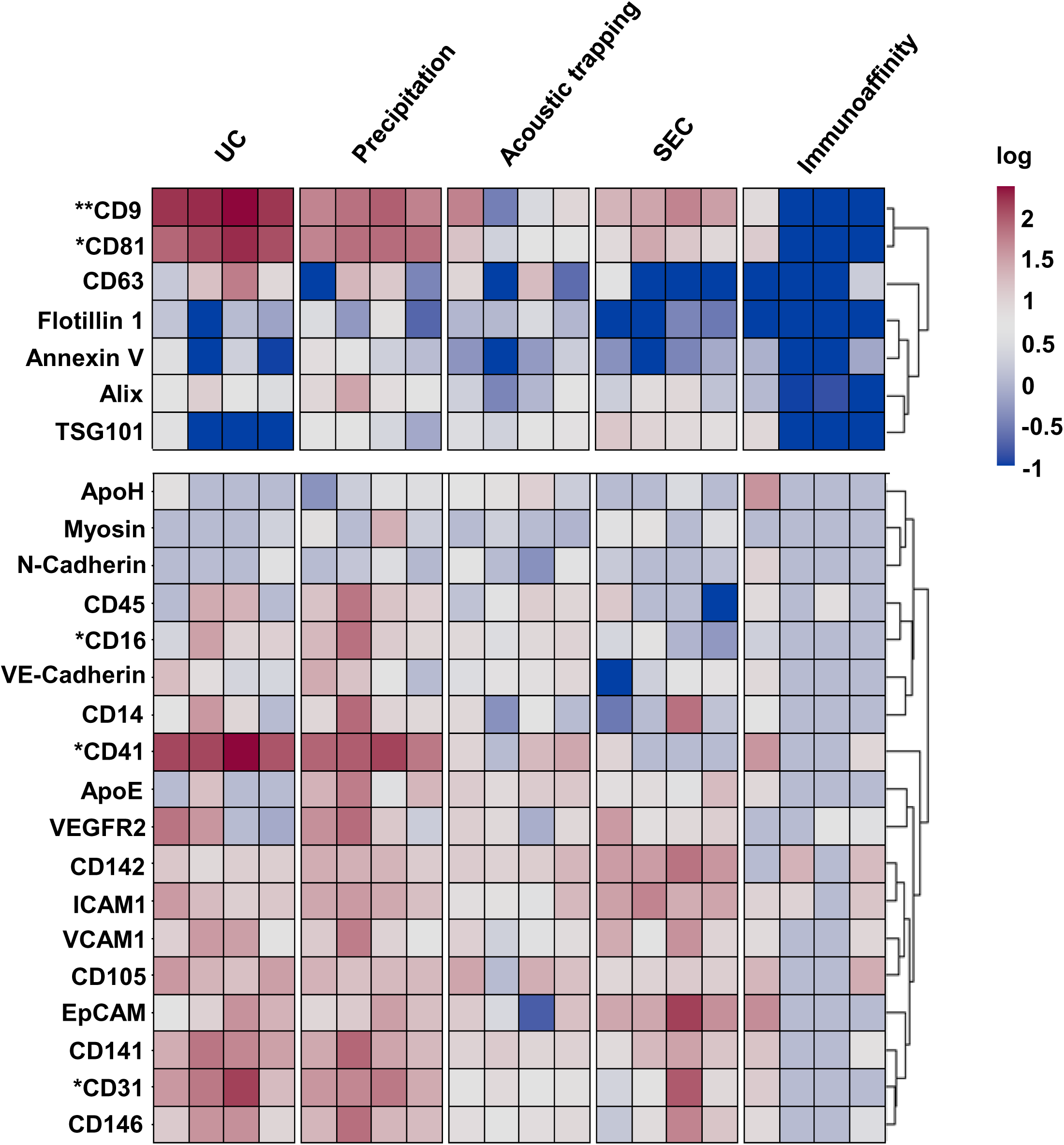
Heatmap of plasma EV derived from different isolation methods using the EV-protein-Array. The heatmap contains extracellular vesicles (EV) markers CD9, CD81, CD63, ALIX, TSG101, Flotillin 1 and Annexin V for ultracentrifugation (UC), precipitation, acoustic trapping, size exclusion chromatography (SEC) and immunoaffinity capture, lipid contaminants apolipoprotein H (ApoH) and apolipoprotein E (ApoE) and cell associated markers. Values are presented as a delta compared to a vehicle control or IgG control and are log normalized (n=4 per isolation method). Data was analyzed by Krusalski-Wallis test with post-hoc Bonferroni correction. *p<0.05, ***p<0.001.

Hierarchical clustering of the EV-Array acquired data for the different isolation methods indicates that there are significant method-dependent differences for cell associated EV markers (**Figure 3**). CD31 was significantly higher in precipitation isolated plasma EV versus acoustic trapping (p<0.05). CD41 was significantly higher in UC isolated plasma EV compared to precipitation isolated EV (p<0.05) and CD16 content was significantly higher in the precipitation isolated plasma EV samples compared to SEC (p<0.01). There were no differences between immunoaffinity capture using CD9, CD63 and CD81 and IgG controls (**Figure 3**).

### Unbiased proteomic analysis of plasma EV

We next determined the proteomic profile of each isolation method using unbiased LC-MS/MS versus their respective PBS or IgG controls for each method. The proteomic profile following plasma EV isolation showed a significantly higher number of quantified protein groups compared to their respective controls for all isolation methods, except immunoaffinity capture, which displayed similar results to the IgG control beads (UC p<0.01, precipitation p<0.05, acoustic trapping p<0.01 and SEC p<0.01, **Figure 4A**). The choice of plasma EV isolation method influenced the overall EV-proteome, but nine protein groups were common across all methods (**Supplemental Figure 2**). These were: immunoglobulin heavy constant gamma 1, alpha-2-macroglobulin, haptoglobin, immunoglobulin heavy constant µ, immunoglobulin kappa constant, serpin family A member 1, albumin, fibrinogen alpha chain and Apo-A1.

**Figure 4:**
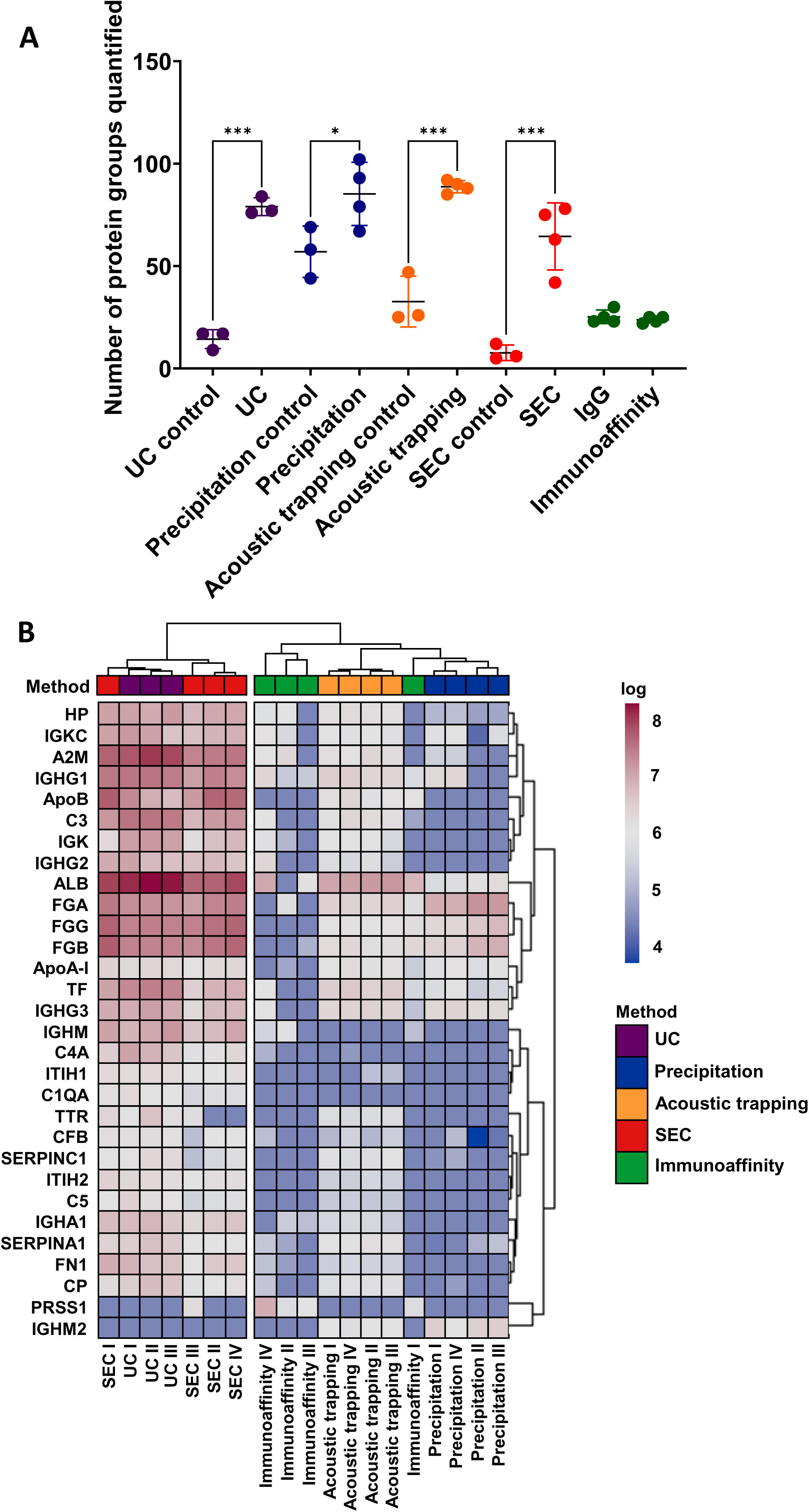
Proteomic comparison of plasma EV isolation methods. **(A)** The number of protein groups quantified by unbiased proteomics for plasma extracellular vesicles (EV) derived by: ultracentrifugation (UC), precipitation, acoustic trapping, size exclusion chromatography (SEC) and immunoaffinity capture, versus control vehicle (PBS) or an IgG control. Protein groups were only included as quantified if they had ≥2 unique peptides. (n=3-4). One-way ANOVA with post-hoc Bonferroni correction. Data are group averages ± standard deviation (SD). *p<0.05, ***p<0.001. **(B)** A heatmap of the top 30 quantified protein groups across all isolation methods. Values from control samples were subtracted to account for background and the values were log normalised. Hierarchical clustering of the isolation methods was conducted using a complete clustering method. (n=3-4).

We used hierarchical clustering of the top 30 quantified protein groups across the different plasma EV isolation methods and found a distinct separation between isolation methods. UC and SEC isolated plasma EV clustered together and precipitation, acoustic trapping and immunoaffinity capture formed a distinct separate cluster (**Figure 4B**). Hierarchical clustering indicates that these differences between UC/SEC and precipitation, acoustic trapping and immunoaffinity capture were driven by an abundance of protein groups with common plasma proteins (ALB, A2M, ApoB, C3, TF, HP, Apo-A1, SERPINA1, CP and ITIH2), fibrinogens (FGG and FGB) and immunoglobulins (IGHG1, IGKC, IGHG3, IGHM, IGK, IGHG2).

To better understand the nature of the proteomic profile for each isolation method, we conducted unbiased Gene Ontology (GO) pathway analysis of the protein groups associated with each isolation method. All methods showed a significant association with EV pathways: Blood Microparticle GO: 0072562 and Extracellular Exosome GO:0070062 (all p<0.01, **Supplemental Figure 3A**). However, there was no clear separation between the different isolation methods using this pathway analysis approach. To further scrutinize the EV proteomic profile obtained per plasma EV isolation method we undertook a statistical comparison using a Fisher’s exact test with published EV-databases EVpedia [31], Vesiclepedia [32] and Exocarta [33]. This determined the similarity between plasma EV proteomic profiles obtained from the five different isolations methods to those published previously by showing the size of the intersect. There was a significant overlap between the five different isolation methods and the archived EV databases (p<0.05, all methods, **Supplemental Figure 3B**). UC, SEC and immunoaffinity capture showed the greatest similarity with published databases. Whereas precipitation and acoustic trapping showed less similarity. Furthermore, we determined whether our plasma EV acquired proteomic profiles from the five different methods were similar to previously published plasma EV proteomic data for Exospin, SEC, ExoQuick, IZON35, IZON70, Optiprep and Exo-easy [16, 17] and found a significant overlap for all five plasma EV isolation methods (p<0.01) (**Supplemental Figure 4)**.

### Targeted sphingolipidomic of plasma EV

EV membranes are largely composed of lipids [46] and lipoproteins are a predominant contaminant in plasma EV samples [41], which are influenced by the choice of isolation method [12, 42]. We compared the lipidomic profile of the different plasma EV isolations methods by undertaking targeted sphingolipidomic analysis. The sphingolipidomic analysis showed a significantly higher number of sphingolipids compared to the respective controls for all isolation methods except immunoaffinity capture (UC p<0.01, precipitation p<0.05, acoustic trapping p<0.01 and SEC p<0.01, **Figure 5A**). However, there were distinct sphingolipidomic differences between the other methods. Eleven of the quantified sphingolipids were common to all methods, which were DhCer(d18:0/24:0), Cer(d18:1/22:0), Cer(d18:1/24:1), Cer(d18:1/24_:0), SM(d18:1/16:0), SM(d18:1/18:0), SM(d18:1/24:1), SM(d18:1/24:0), HexCer(d18:1/16:0), HexCer(d18:1/24:1) and LacCer(d18:1/24:1) (**Supplemental Figure 5**). Hierarchical clustering analysis of the sphingolipidomic profile for each of the isolation methods showed that there are three distinct clusters (**Figure 5B**). Immunoaffinity capture and precipitation formed two separate individual clusters, whilst UC, acoustic trapping and SEC clustered together. Hierarchical clustering indicates that these group differences were driven by an abundance of sphingomyelins (16:0, 18:0, 24:0 and 24:1), ceramides (22:0, 24:0 and 24:1), hexosylceramides (24:1) and lactosylceramides (16:0) in the precipitation group and a lack of these sphingolipids in the immunoaffinity capture group.

**Figure 5:**
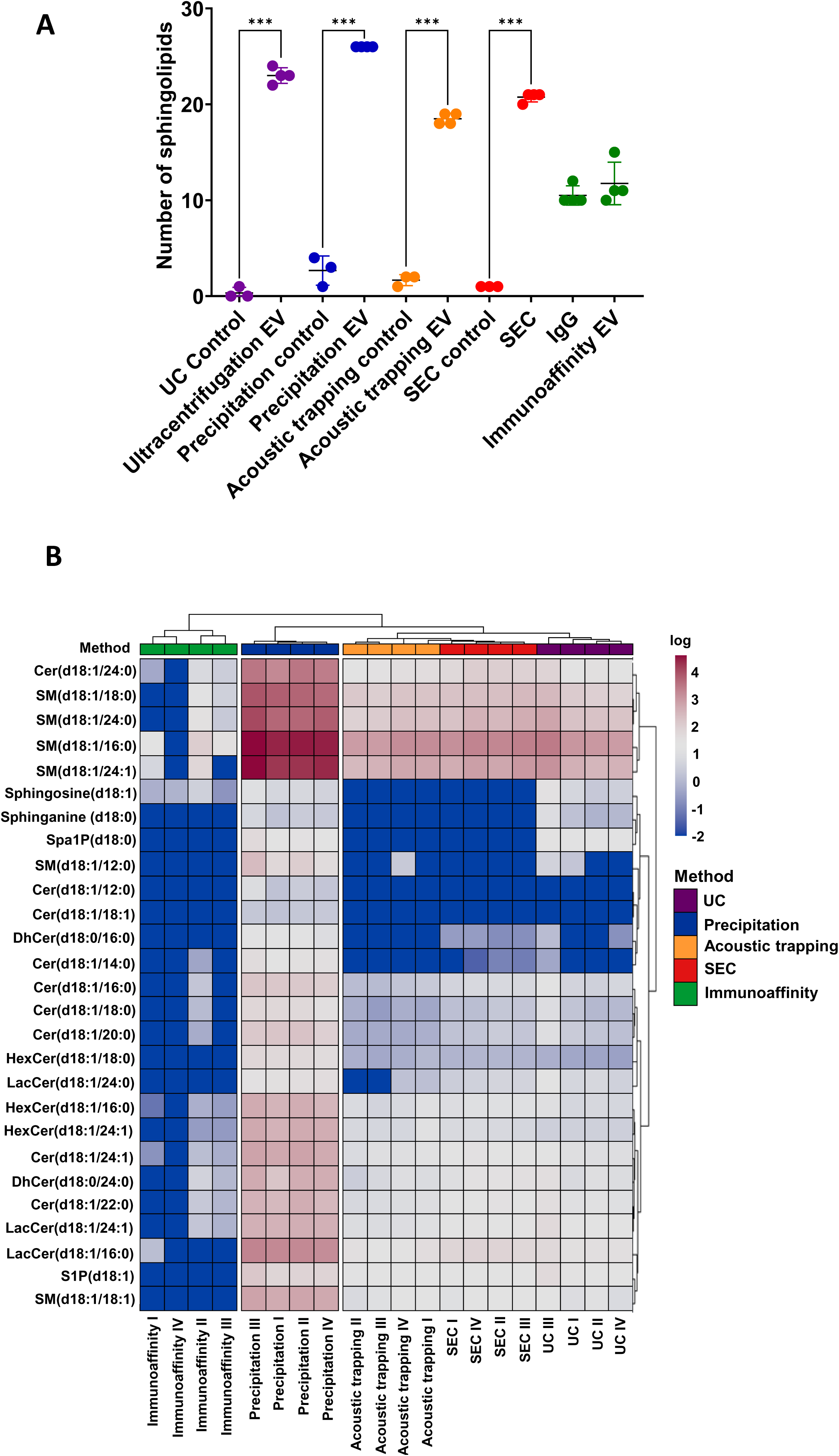
Sphingolipidomic analysis of plasma EV from different isolation methods. **(A)** Number of sphingolipids quantified in plasma extracellular vesicles (EV) isolated by ultracentrifugation (UC), precipitation, acoustic trapping, size-exclusion chromatography (SEC) and immunoaffinity capture and subjected to targeted sphingolipid analysis versus control vehicle (PBS) or an IgG control. Data are group averages ± standard deviation (SD) and were analysed by One-way ANOVA with post-hoc Bonferroni correction. ***p<0.001. (n=4) **(B)** Heat map of plasma EV sphingolipids. Values from control samples were subtracted to account for background and the values were log normalised. Hierarchical clustering of the isolation methods was conducted using a complete clustering method. (n=3-4). Data were analysed by Krusalski-Wallis test with post-hoc Bonferroni correction.

### Integrated comparison of plasma EV isolation methods

These data shows that plasma EV isolation methods have divergent impact on the yield of EV, their purity, the type of EV isolated from plasma that are associated with a particular cell source and the overall proteomic and sphingolipidomic profile. To better understand how these individual method associated differences influenced the plasma profile, we undertook integrated analysis of all the different plasma EV isolation methods with all of the acquired data. This included the EV particle concentration, protein concentration, EV-protein array data, proteomic and sphingolipidomic data. To condense the multiple different variables, we converted the data into principal components. Principal component analysis with two components (PCA1 and PCA2) accounted for 47.2% of the variance and showed clear separation between the different isolation methods (**Figure 6A/B).** UC, SEC and precipitation form distinct clusters, whereas acoustic trapping and immunoaffinity capture clustered together. Specific principal component analysis indicated that the PCA1 is driven by proteomics acquired data and PCA2 is driven by sphingolipidomic data (**Supplementary Figure 6**). These integrated data show that the EV isolation method influences the omic and integrated-based plasma EV-profile, which may influence interpretation of EV-acquired data for clinical biomarker discovery and precision diagnostics.

**Figure 6:**
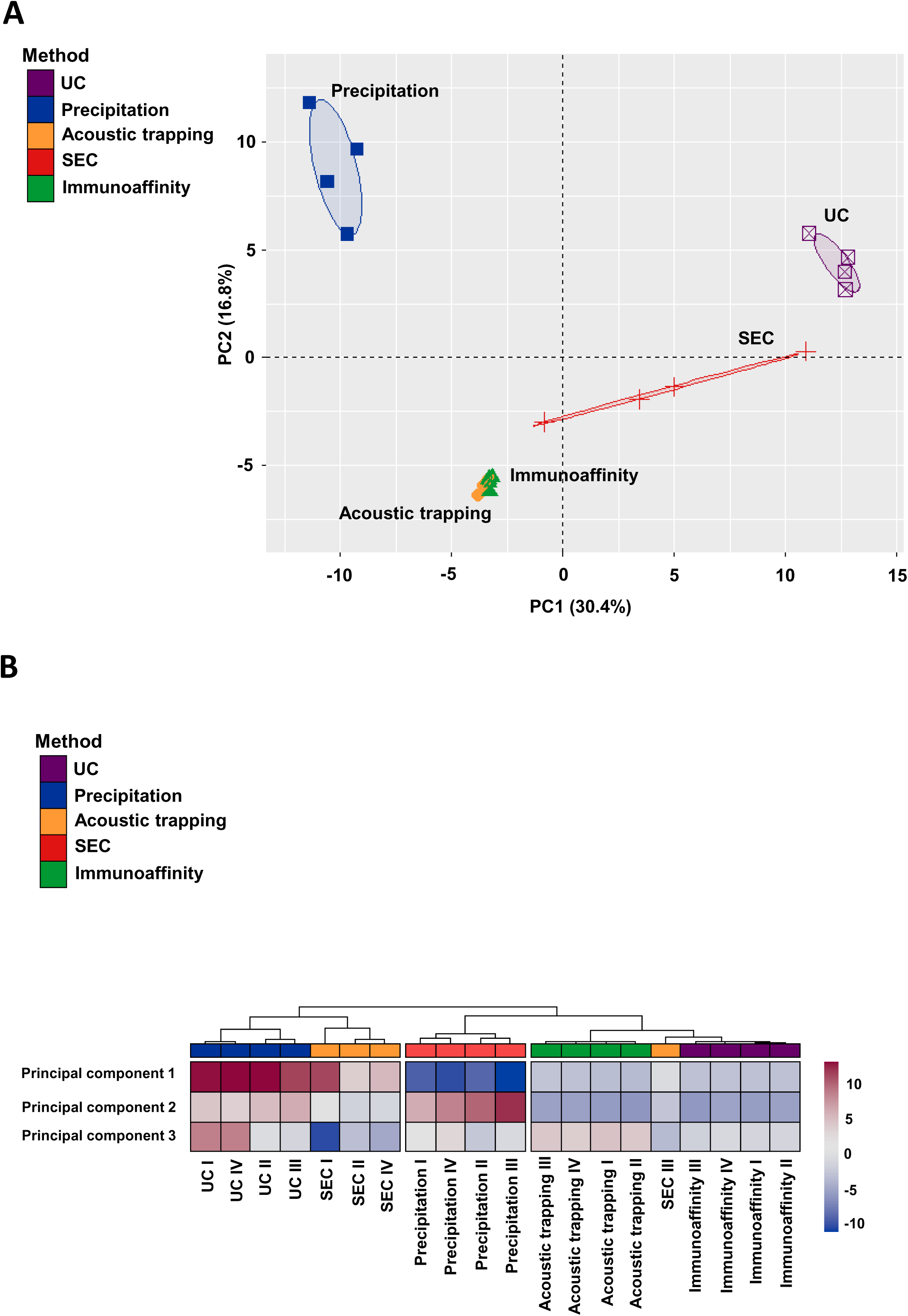
Principal component analysis of plasma EV characteristics following data integration. **(A)** A principal component (PC) analysis of plasma extracellular vesicles (EV) isolated by ultracentrifugation (UC), precipitation, acoustic trapping, size-exclusion chromatography (SEC) and immunoaffinity capture: including plasma EV concentration, protein concentration, EV-protein-Array, proteomics and sphingolipidomics. The integrated data was condensed to PC1 and PC2. **(B)** A heatmap with the various principal components to compare the different isolation plasma EV isolation methods. Hierarchical clustering of the isolation methods was conducted using a complete clustering method. (n=3-4).

### Plasma EV isolation methods influence the diagnostic potential of plasma EV from patients following MI

We next determined whether the choice of plasma EV isolation method impacts the ability to detect changes in EV-profile in a disease state. We obtained plasma at time of presentation with MI, but prior to percutaneous coronary intervention (PCI), and a matched control plasma sample was obtained at 1-month post-MI from the same patients. The clinical patient characteristics are detailed in **Table 2**.

**Table 2:** Clinical characteristics of the MI patients. Age, sex (M/F), glucose, white blood cells counts, troponin peak, cholesterol, diabetes status, smoker status, infarct size determined by late gadolinium enhancement MRI 6 months post-AMI and left ventricle ejection fraction (LVEF %) 6 moths post-MI. Data are group averages ± standard deviation (SD) (n=6).

|  | Average ± SD |
| --- | --- |
| Age | 67.0 ± 11.4 |
| Sex (male/female) | 6/0 |
| Glucose (mmol/L) | 5.7 ± 0.5 (N=3) |
| White blood cell count (x10^9) | 7.6 ± 2.2 |
| Troponin (peak ng/L) at presentation | 358.7 ± 401.2 |
| Cholesterol (mmol/L) | 4.8 ± 1.2 |
| Diabetic status (diabetes / non) | 0/6 |
| Smoker status (smoker / non) | 1/5 |
| Infarct size at 6-months | 15 ± 7.8 |
| LVEF at follow up (%) | 47 ± 5.0 |

### Plasma EV concentration in MI is influenced by the isolation method

To mitigate any potential variability in bio-banked plasma samples we used three technical plasma replicates per patient, per time point and per method. There were significantly more plasma EV at time of presentation with MI versus the 1 month control follow up when plasma EV were isolated by UC, precipitation and acoustic trapping (p<0.05 for all) (**Figure 7A**), but not by SEC and immunoaffinity capture (UC presentation: 2.8 x 10^9^ EV / mL vs. UC follow-up: 2.1 x 10^9^ EV / mL; precipitation presentation: 1.5 x 10^12^ EV / mL vs. precipitation follow-up: 7.9 x 10^11^ EV / mL; acoustic trapping presentation: 4.3 x 10^10^ EV / mL vs. acoustic trapping follow-up: 1.9 x 10^10^ EV / mL; SEC presentation: 2.5 x 10^11^ EV / mL vs. SEC follow-up: 9.2 x 10^10^ EV / mL and immunoaffinity presentation: 2.0 x 10^11^ EV / mL vs. immunoaffinity capture follow-up: 9.8 x 10^10^ EV / mL, **Figure 7A**). The technical variance between the three independent isolations from each patient, and at each time, point showed that SEC gave significantly less variance compared to UC (10.0 ± 3.1 % vs. 22.7 ± 13.0 %, p<0.01) (**Supplemental Figure 7A**). The mean plasma EV size was similar between the time of presentation and 1 month follow up control for the five different plasma EV isolation methods (**Supplemental Figure 7B**) and the size and concentration distribution profile showed no distinct differences between time points or dependent on the plasma EV isolation method (**Supplemental Figure 7C**).

**Figure 7:**
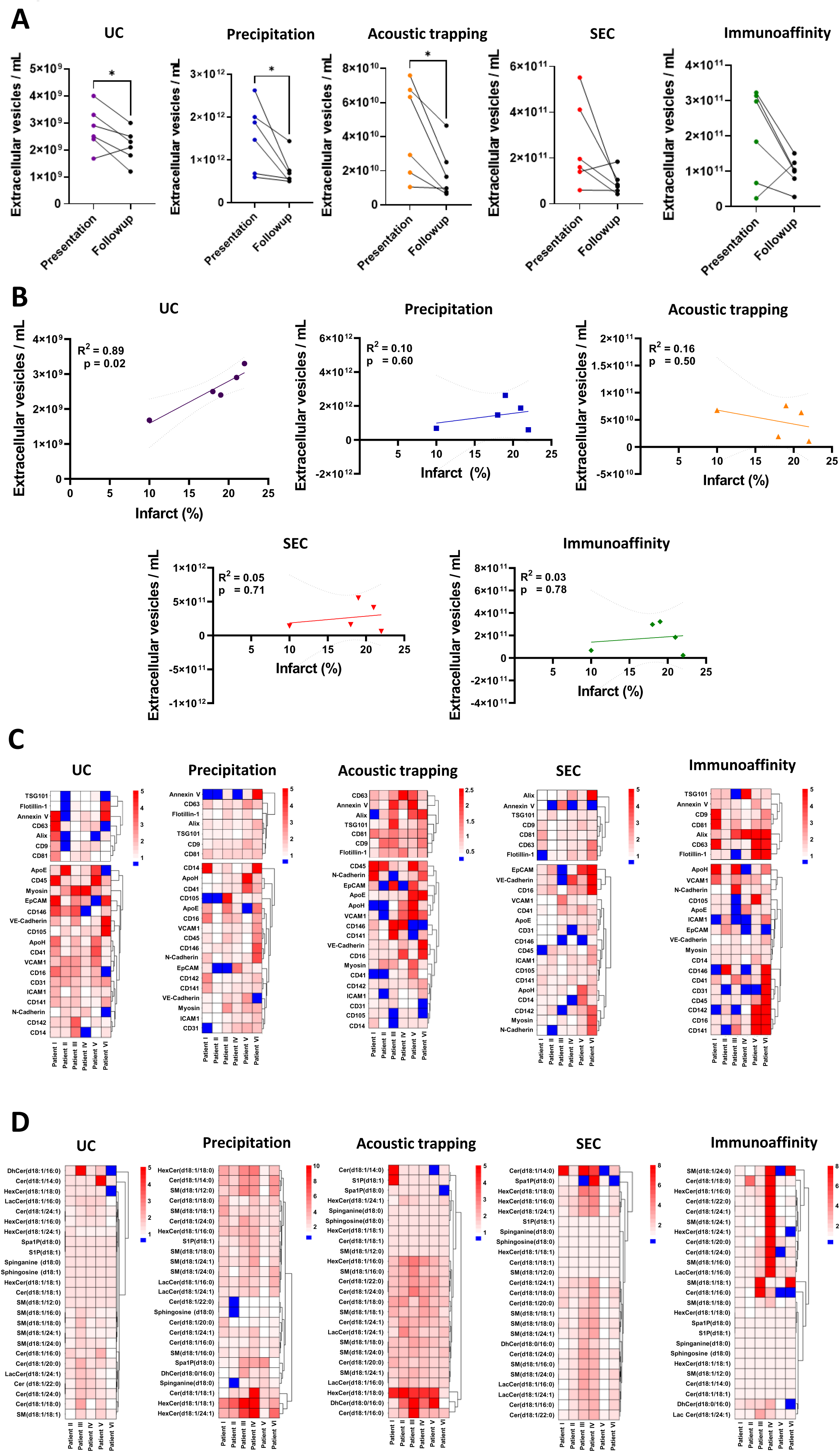
Plasma EV analysis using different isolation methods in patients following presentation with myocardial (MI) infarction compared to samples from the same patients after a 1-month follow up. **(A)** Comparison of concentration and size-distribution profile by Nanoparticle Tracking Analysis of plasma extracellular vesicles (EV) from patients following presentation with MI and after a 1-month follow up using: ultracentrifugation (UC), precipitation, acoustic trapping, size exclusion chromatography (SEC) and immunoaffinity capture (n=6 per timepoint). Data are group average ± standard deviation (SD). Paired T-test analysis *p<0.05. **(B)** A Pearson correlation analysis of plasma EV / mL derived from each isolation method at time of presentation vs. the infarct size determined by cardiac MRI using late gadolinium enhancement 6-months post-infarct (n=5). **(C)** Heatmap of plasma EV-Array analysis. The top section of the heatmap contains the different EV markers CD9, CD81, CD63, ALIX, TSG101, Flotillin 1 and Annexin V. Values are presented as fold over the respective matched follow up control sample. **(D)** Heatmap showing the plasma EV sphingolipidomic profiles in MI patents at presentation versus a 1-month follow up for UC, precipitation, acoustic trapping, SEC and immunoaffinity capture. Values are presented as fold over the respective matched follow up.

### The plasma EV isolation method influences plasma EV concentration association with infarct size

We have previously reported that the total concentration of plasma EV in the peripheral blood isolated by UC at the time of presentation correlates with the size of myocardial injury and scar, determined by late gadolinium enhanced (LGE) MRI 6 months post-MI [5, 6]. Therefore, we determined whether plasma EV concentrations from the five different plasma EV isolation methods influenced the ability to determine this important clinical association. Infarct size at 6-months post-MI significantly correlated with the plasma EV concentration at time of presentation for UC samples (R^2^ = 0.89, p=0.02), but not for precipitation, acoustic trapping, SEC or immunoaffinity capture (R^2^=0.10 p=0.60, R^2^=0.16 p=0.50, R^2^=0.05 p=0.71 and R^2^=0.10, p=0.78, respectively) (**Figure 7B**).

### Cell associated plasma EV are not influenced by the isolation method in MI

We determined whether the isolation methods influenced cell associated plasma EV-markers in MI patients at presentation versus the respective 1 month follow up sample. The high through put EV-Array data was expressed as fold change over the matched follow-up control samples per patient. Plasma EV markers were increased at time of presentation vs. follow-up control (**Figure 7C**). However, the pattern of change in plasma EV-markers were not consistent across the different isolation methods (**Figure 7C**). Hierarchical clustering of the plasma EV marker response at the time of presentation with MI showed method-dependent clustering. Similarly, cell associated markers on plasma EV were differentially enriched following MI (**Figure 7C**). Once again, unbiased clustering showed method-dependent groupings. Together these data suggest that changes in plasma-EV protein are influenced by the choice of plasma EV isolation method following MI in isolation and when clustered together.

### Plasma EV sphingolipidomic profiles are altered following MI and influenced by the isolation method

Next, we profiled EV-sphingolipids in the acute phase following MI (calculated as fold over matched follow-up control samples) using the different plasma EV isolation methods. Plasma EV-sphingolipidomic profiles for samples generated by precipitation, acoustic trapping and SEC contained significantly more quantified lipid groups at time of presentation with MI versus samples obtained by UC and immunoaffinity capture (p<0.01, all). We ranked plasma EV-lipid profiles based on their overall abundance of lipid groups and found that ceramides were significantly higher in the precipitation isolated plasma EV samples, followed by SEC, acoustic trapping, ultracentrifugation / immunoaffinity capture (p<0.01). Whereas the sphingomyelins were significantly higher in the precipitation group versus all other isolation methods (p<0.001) (**Figure 7D**). The clustered plasma EV-sphingolipid profile following MI was influenced by the choice of isolation method. Several ceramides and sphingomyelins were significantly higher in the plasma EV isolated by precipitation or acoustic trapping compared to UC (**Supplemental Figure 8**). In addition, the fold increase of Cer(d18:1/22:0), SM(d18:1/18:1) and sphinganine (d18:0) in plasma EV isolated by precipitation at time of presentation vs. 1-month follow up significantly correlated with the infarct size of the patients at 6-months post-MI (R^2^ = 0.91 p=0.01, R^2^ = 0.78 p=0.05 and R^2^ = 0.78 p=0.05, respectively) (**Supplemental Figure 9**). This result further confirmed that plasma EV isolation methods, using targeted sphingolipidomic methods, are associated with unique method dependent profiles in patients following MI.

### Integrated-omics comparison of plasma EV isolation methods in MI patients

Finally, we integrated all the acquired plasma EV data from the MI patients in the acute presentation and follow-up time points. A principal component analysis was constructed to assess if the plasma EV profile at time of presentation differed from follow-up using a 95% confidence level. Integrated analysis showed that plasma EV profiles were only distinguishable at time of presentation with MI for precipitation and acoustic trapping, but not for those plasma EV isolated by UC, SEC and immunoaffinity capture (**Figure 8**). These findings indicate that different plasma EV isolation methods influence the diagnostic potential of plasma liberated EV following MI when multiple datasets are integrated to determine plasma EV profiles for potential panel biomarker discovery.

**Figure 8:**
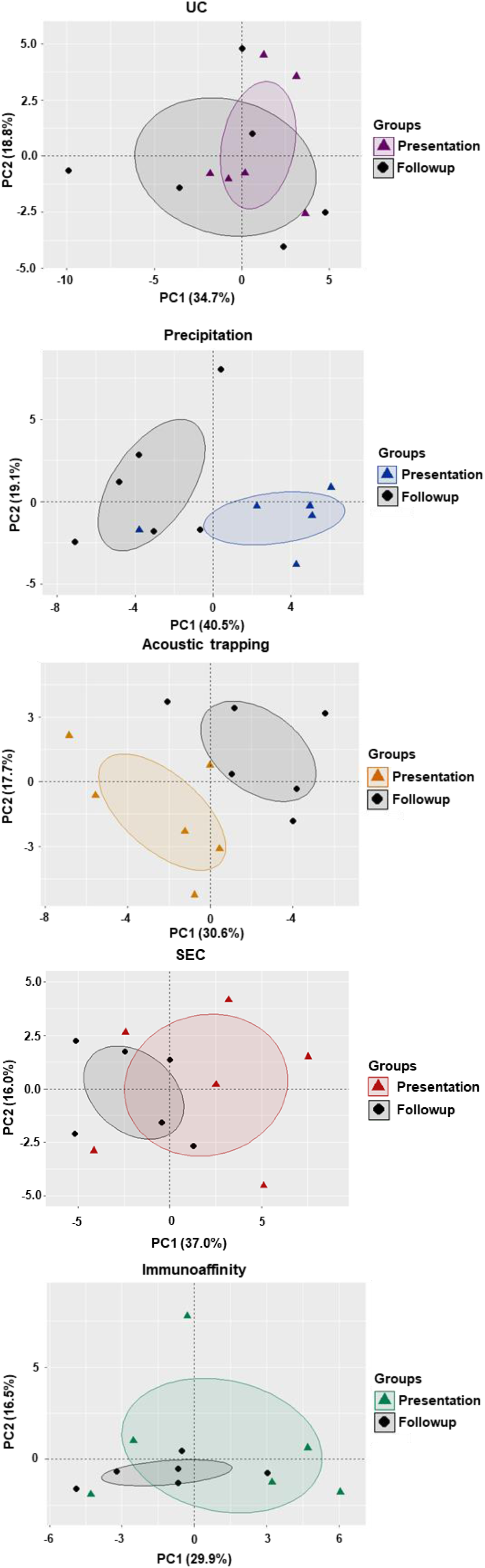
Principal component analysis of integrated plasma EV characterisation at time of presentation with myocardial infarction (MI) vs. 1-moth follow-up in the same patients using different plasma EV isolation methods. A principal component (PC) analysis of plasma EV isolated by ultracentrifugation (UC), precipitation, acoustic trapping, size-exclusion chromatography (SEC) and immunoaffinity capture from patients presenting with MI versus a 1-month follow up control. Plasma EV characteristics include including plasma EV concentration, protein concentration, EV-protein-Array and sphingolipidomics. The integrated data was condensed to PC1 and PC2. (n=5-6). The eclipses indicate the 95% confidence interval.

## Discussion

Plasma EV-cargo may provide unparalleled insight into tissue homeostasis and pathological processes to facilitate identification of patients for focused therapies, but the methods to isolate plasma EV may impact the EV characteristics. Here, we found that; (I) the choice of plasma EV isolation method affected the plasma EV concentration, sphingolipid and proteomic profile, (II) but the five different plasma EV isolation methods shared a common EV protein and sphingolipid profile. (III) Plasma EV isolation by immunoaffinity capture using anti-CD9, -CD63 and -CD81 coated antibody beads with the current protocol yields a similar profile to IgG control beads. (IV) The isolation method affected the ability to detect alterations in plasma EV sphingolipids and proteins in MI patients and (V) their association with infarct size determined by cardiac MRI 6 months post-MI. For example, the plasma EV concentration at time of presentation obtained using UC provides prognostic information, but this method is less suitable as a tool to generate EV with higher purity for use in mechanistic studies.

Previous studies have sought to determine the influence of plasma EV isolation methods, but have often focused on comparing the net influence on the EV-proteome [3, 16, 17, 43] or RNA profiles [16] after isolation. These approaches can neglect the increased dimensionality of EV components, which carry numerous biologically active molecules such as proteins, lipids, RNA/DNA and metabolites. Furthermore, there is varied assessment of contaminations in EV preparations for use in ‘omics’ studies, which may mask or skew the interpretation of datasets. The approach employed here utilizes the same independent biological replicates across multiple methods for plasma EV isolation and characterization, in conjunction with a control PBS/vehicle sample or an IgG for immunoaffinity capture beads. Here, the repeated analysis of the same plasma samples across multiple methods has enabled individual biological variability and potential plasma EV isolation irregularities to be explored. By using individual and integrated analysis, we showed that each method of plasma EV isolation was internally consistent; in so much that the samples clustered according to their isolation method in unsupervised hierarchical clustering and principal component analysis of all the acquired data.

Consistent with previous reports, our plasma EV-proteome was dependent on the plasma EV isolation method [16, 17]. However, our data shows that there are common plasma proteins, such as albumin and apolipoproteins ApoA-I and ApoB, present in all five isolation methods tested, with significant similarities to archived EV-databases [31–33] and published studies [16, 17, 31]. In particular, the plasma EV-proteome indicated that UC/SEC cluster together based on the top 30 protein groups quantified, due to higher quantities of fibrinogens and immunoglobulins compared to the other methods. This is, potentially, a methodological constraint of employing untargeted mass spectrometry on plasma EV, which favors the most abundant peptides.

To better determine the protein profile of known plasma EV markers and cell associated markers, we utilized a high throughput EV-Array [50] following plasma EV isolation from the five different methods. We found that CD9 and CD81 were the most abundant in plasma EV derived by UC versus precipitation and SEC in healthy volunteers. Additionally, we found that cell-associated markers such as CD41 (platelets) and CD16 (immune cells) were enriched in UC and precipitation compared to other methods only in healthy volunteers. This high throughput antibody EV-Array circumvents issues of non-EV associated protein and lipid contaminants in plasma EV preparations. However, the five different plasma EV isolation methods used here may have influenced the EV-protein corona. Proteins such as albumin and apolipoproteins (ApoA-I, ApoB, ApoC-III and ApoE) show an association with the surface of EV [45], which may influence antibody mediated binding to the array or subsequent detection of EV-associated proteins in this sandwich ELISA like technique. Protein coronas found on EV are affected by the isolation method [45] and our data shows that the plasma EV-proteome and EV-Array acquired data are also influenced by the isolation method. Indeed, our western blot and proteomic data reported considerable variation in the amount of albumin across the methods, which is a predominant protein in the EV protein corona. In particular, precipitation isolated preparations was associated with lower albumin, which could be due to the presence of PEG and high salt concentrations in the isolation buffer that might be expected to influence albumin associations with the EV protein corona. The temporal dynamics of the EV-protein and possibly EV-lipid corona are poorly understood, and further elucidation of these interactions will be essential to understand the data from these preparations.

Immunoaffinity based methods, such as magnetic and polystyrene bead conjugations to specific antibodies, such as tetraspanins (CD9, CD63 and CD81), have been heralded as an important advancement in the capture of highly pure EV populations from the plasma. Immunoaffinity based methods could circumvent issues of lipoprotein and protein contamination in plasma EV yields, which are common in methods that use EV physical characteristics such as size and density for isolation such as UC and SEC [16, 46]. To the best of our knowledge this is the first study to compare magnetic beads coated with antibodies for tetraspanins (CD9, CD63 and CD81) with a matched IgG control for plasma EV isolation, using detailed integrated analyses. The plasma EV-profile of proteins and lipids for the immunoaffinity beads and matched IgG control was similar by Western blot, EV-protein-array, the number of peptides groups in proteomic profiles, GO pathway analysis and targeted sphingolipidomics. TEM imaging of IgG beads showed no visible EV-like particles or membranous structures, indicative of whole or sheared EV-particles, captured to the bead surface. However, we hypothesize that soluble non-EV-associated proteins (such as C3, A2M, ALB and fibrinogens A and B) and lipids (such as ApoB, ApoE and ApoA-I) are interacting with IgG beads, which have become associated with EV-profiles in omics studies and in archived databases [31–33]. Studies using less complicated matrices, like conditioned cell culture media, show a more robust distinction between anti-CD9, CD63 and CD81 beads versus IgG control beads [8, 47], indicating that the similarity between tetraspanin and IgG antibody beads here might be matrix specific. Future plasma EV studies using antibody-bead isolation should include a matched IgG control per characterisation when using omic-approaches

Having determined plasma EV isolations method similarities and differences on the EV-characteristics and cargo in healthy volunteers, we assessed the influence of plasma EV isolation methods in a clinical situation where plasma EV number and composition are altered. We have previously shown that plasma EV are altered following MI to mediate long range signalling and induce the mobilization of splenic-neutrophils, splenic-monocytes and orchestrate their transcriptional programming [5, 6]. The choice of plasma EV isolation method determined whether there was a higher concentration of EV immediately following MI compared to a 1-month follow-up control sample from the same patients. The new data here support our previous observations that the concentration of plasma EV isolated by UC from the time of presentation with MI correlates with the infarct size [5]. However, plasma EV concentrations derived from the other isolation methods in the same patients did not associate with infarct size. One possible explanation is the modest sample size utilised in this multi-method comparison (N=6), which is smaller than those utilised by us in previous publications (N=15-22) [5, 6].

Plasma EV sphingolipids are predictive of MI [3]. However, the influence of plasma EV isolation methods on the plasma EV-sphingolipidomic profile were not reported. We found a distinct sphingolipid profile between the plasma EV isolation methods. Precipitation-based EV isolation yields a significantly higher lipid content, principally due to higher proportions of sphingomyelins and ceramides. Sphingomyelins are found in cell membranes and associated with high- and low-density lipoproteins [48, 49]. The high presence of sphingomyelins and ceramide in the plasma EV profile of the precipitation-based method is likely due to the co-isolation of lipoproteins. PEG-based solutions co-isolate lipoproteins [56]. Lipoproteins contain ApoA-I and ApoB, which were higher in plasma EV derived by precipitation-based isolation when compared to the other methods by Western blot.

Lipoproteins can also masquerade as EV-like particles in dynamic light scattering in techniques such as NTA [12]. Precipitation isolated plasma EV had the highest concentration of particles / mL when compared to other methods. EV Cer(d18:1/20:0) only showed elevation in precipitation-based isolation compared other methods. Similarly, precipitation isolated EV showed sphinganine(d18:0) correlated with infarct size at 6-months, but this was not the case for the other methods. Analysis of plasma EV-sphingolipid remains challenging due to constraints due to lipoparticle remnants [57]; however we show here for the first time there are common sphingolipids for different plasma EV isolation methods. In particular, plasma EV-protein and EV-sphingolipids clustered uniquely based on the isolation method, which may impact plasma EV diagnostics where a panel of proteins and sphingolipids are used for differentiation of clinical disease or outcome.

In summary, our data show that the choice of plasma EV isolation method influences the concentration of plasma EV, the EV-proteome and EV-sphingolipid profile in healthy volunteers and MI patients, where methodological differences determined associations with infarct size. Precipitation based plasma EV isolation gives the highest particle concentration and enriches for more sphingolipids, but co-isolate large quantities of apolipoproteins. In addition, immunoaffinity capture by using antibodies against tetraspanins yields similar EV-protein and EV-sphingolipid profiles to IgG control beads. Unbiased integrated analysis shows that plasma EV isolation methods cluster independently, but despite the selection predispositions imposed by each method, a core of EV associated proteins and lipids was detectable using all the approaches.

## Acknowledgments

DP acknowledges his funding from the Department of Pharmacology, the Alison Brading Memorial Fund Lady Margaret Hall, the Clarendon Fund provided by the University of Oxford and the Medical Research Council (MR/N013468/1). NA and RC acknowledge support by research grants from the British Heart Foundation (BHF) Centre of Research Excellence, Oxford (NA and RC: RE/13/1/30181 and RE/18/3/34214); British Heart Foundation Project Grant (NA and RC: PG/18/53/33895); the Tripartite Immunometabolism Consortium, Novo Nordisk Foundation (RC: NNF15CC0018486); Oxford Biomedical Research Centre (BRC); Nuffield Benefaction for Medicine and the Wellcome Institutional Strategic Support Fund (ISSF) (NA) and a Health Research Bridging Salary Scheme (HRBSS) to N.A. BZ work was supported by funding from the European Union’s Horizon 2020 Research and Innovation Program under Marie Sklodowska-Curie Grant Agreement 812890, ArthritisHeal. The views expressed are those of the author(s) and not necessarily those of the National Health Service, the National Institutes of Health Research or the Department of Health

## Declaration statement

The Author(s) declare(s) that there is no conflict of interest.

## Supplemental Figures

**Supplemental Figure 1: The ratio of plasma extracellular vesicles (EV) to protein concentration per isolation method:** ultracentrifugation (UC), precipitation, acoustic trapping, size-exclusion chromatography (SEC) and immunoaffinity capture. EV are expressed as per mL and protein concentration is expressed µg. Values are expressed as delta over phosphate buffer solution (PBS) control or IgG control. Data are group averages ± standard deviation (SD) (n=4). One-way ANOVA with post-hoc Bonferroni correction. **p<0.01, ***p<0.001.

**Supplemental Figure 2: Venn diagram of quantified protein groups in different plasma extracellular vesicle (EV) isolation methods:** ultracentrifugation (UC), precipitation, acoustic trapping, size-exclusion chromatography (SEC) and immunoaffinity capture. Quantified protein groups within a method were pooled after subtraction of phosphate buffer solution (PBS) control or IgG control. Protein groups that had more than 1 repeat within a method were included in the Venn diagram.

**Supplemental Figure 3: Gene Ontology analysis of plasma extracellular vesicle (EV) isolation method proteomics:** ultracentrifugation (UC), precipitation, acoustic trapping, size-exclusion chromatography (SEC) and immunoaffinity capture. **(A)** Quantified protein groups were expressed as delta over phosphate buffer solution (PBS) control or IgG control. Quantified protein groups that appeared more than once per method were included in the Gene Ontology (GO) analysis. The GO cellular components associated with each method were ranked based on the false discovery rate (FDR) and the top five were included in the tables. In addition, the GO enrichment scores were plotted against the false discovery rate p-values in the scatterplot. **(B)** The overlap between quantified protein groups per plasma EV isolation method and published EV proteomics databases EVpedia, Exocarta and Vesiclepedia. The table contains the percentage of overlapping protein groups vs. total protein groups per isolation method, with a p-value calculated by fisher-exact test.

**Supplemental Figure 4: The overlap between the number of quantified protein groups per plasma extracellular vesicle** (**EV) isolation method and published plasma EV proteomic datasets.** Quantified protein groups within a method were pooled after subtraction of phosphate buffer solution (PBS) control or IgG control. Protein groups that had more than 1 repeat within a method were included. Plasma EV-proteomes determined by ultracentrifugation (UC), precipitation, acoustic trapping, size-exclusion chromatography (SEC) and immunoaffinity capture were compared with published EV proteomic datasets. The overlap of protein groups between each method were compared and listed out of the total protein groups quantified.

**Supplemental Figure 5: Venn diagram showing the overlap between quantified sphingolipids in different plasma extracellular vesicles (EV) isolation methods:** ultracentrifugation (UC), precipitation, acoustic trapping, size-exclusion chromatography (SEC) and immunoaffinity capture. Sphingolipids within a method were pooled after subtraction of phosphate buffer solution (PBS) control or IgG control. Sphingolipids that had more than 1 repeat within a method were included in the Venn diagram.

**Supplemental Figure 6: Scree plots of each principal component following the integrated data analysis of plasma extracellular vesicle (EV) isolated with the different methods:** ultracentrifugation (UC), precipitation, acoustic trapping, size-exclusion chromatography (SEC) and immunoaffinity capture. Data from the acquired unbiased proteomics, targeted sphingolipidomics, EV-Array analysis and concentration/size analysis was pooled for each isolation method. Principal components were generated with the R-package *Factoextra* and *FactoMineR.* Each scree plot indicates the percentage of variance explained for the principal component and the top 30 drivers for the principal component. Data was labelled based on the origin; Lipid indicates data from the targeted sphingolipid data, EV indicates data from the EV-protein-array and unlabeled was selected for data from the proteomics.

**Supplemental Figure 7: Plasma extracellular vesicle (EV) concentration and size characteristics after isolation from myocardial infarction (MI) patients at time of presentation and one month follow up. (A)** The variance of plasma EV concentration isolated by each method measured by Nanoparticle Tracking Analysis (NTA). The variance was calculated by relative standard deviation over three different measurements. Values are expressed as delta over phosphate buffer solution (PBS) control or IgG control. **(B)** The average size of the isolated plasma EV for each method at time of presentation vs. follow up measured by NTA. Immunoaffinity capture can not be acquired. **(C)** Size distribution profiles of the plasma EV from MI patients at time of presentation and one month follow up. Immunoaffinity capture can not be acquired. Data are group averages ± standard deviation (SD) (n=12 for **A**, n=6 for **B,** n=6 per time point for **C**). One-way ANOVA with post-hoc Bonferroni correction. *p<0.05 for **A** and **B**.

**Supplemental Figure 8: Sphingolipid concentrations of plasma extracellular vesicles (EV) isolated by different methods:** ultracentrifugation (UC), precipitation, acoustic trapping, size-exclusion chromatography (SEC) and immunoaffinity capture. The fold change in sphingolipid concentration in plasma EV isolated at time of presentation with myocardial infarction (MI) versus the concentration in plasma EV isolated 1-month post-MI per isolation method. Data are group averages ± standard deviation (SD) and were analysed by one-way ANOVA with post-hoc Bonferroni correction. *p<0.05 and **p<0.01. (n=5-6)

**Supplemental Figure 9: Plasma extracellular vesicle (EV) sphingolipid concentration correlation with infarct size.** A Pearson correlation analysis was conducted with the the fold change in sphingolipid (**A**: Cer(d18:0/22:0); **B**: sphinganine (d18:0); **C**: SM(d18:1/18:1) concentration in plasma EV isolated at time of presentation with myocardial infarction (MI) versus the concentration in plasma EV isolated 1-month post-MI by the precipitation method versus the infarct size determined by cardiac MRI using late gadolinium enhancement 6-months post-infarct (n=5).

## References

1. de Miguel Pérez, D., et al., Extracellular vesicle-miRNAs as liquid biopsy biomarkers for disease identification and prognosis in metastatic colorectal cancer patients. Sci Rep, 2020. 10(1): p. 3974.

2. Dong, L., et al., Circulating Long RNAs in Serum Extracellular Vesicles: Their Characterization and Potential Application as Biomarkers for Diagnosis of Colorectal Cancer. Cancer Epidemiol Biomarkers Prev, 2016. 25(7): p. 1158–66.

3. Burrello, J., et al., Sphingolipid composition of circulating extracellular vesicles after myocardial ischemia. Sci Rep, 2020. 10(1): p. 16182.

4. Kalani, M.Y.S., et al., Extracellular microRNAs in blood differentiate between ischaemic and haemorrhagic stroke subtypes. J Extracell Vesicles, 2020. 9(1): p. 1713540.

5. Akbar, N., et al., Endothelium-derived extracellular vesicles promote splenic monocyte mobilization in myocardial infarction. JCI Insight, 2017. 2(17).

6. Akbar, N., et al., Rapid neutrophil mobilisation by VCAM-1+ endothelial extracellular vesicles. Cardiovasc Res, 2022.

7. Evander, M., et al., Non-contact acoustic capture of microparticles from small plasma volumes. Lab Chip, 2015. 15(12): p. 2588–96.

8. Kowal, J., et al., Proteomic comparison defines novel markers to characterize heterogeneous populations of extracellular vesicle subtypes. Proc Natl Acad Sci U S A, 2016. 113(8): p. E968–77.

9. Onódi, Z., et al., Isolation of High-Purity Extracellular Vesicles by the Combination of Iodixanol Density Gradient Ultracentrifugation and Bind-Elute Chromatography From Blood Plasma. Front Physiol, 2018. 9: p. 1479.

10. Nordin, J.Z., et al., Ultrafiltration with size-exclusion liquid chromatography for high yield isolation of extracellular vesicles preserving intact biophysical and functional properties. Nanomedicine, 2015. 11(4): p. 879–83.

11. Geyer, P.E., et al., Plasma Proteome Profiling to detect and avoid sample-related biases in biomarker studies. EMBO Mol Med, 2019. 11(11): p. e10427.

12. Brennan, K., et al., A comparison of methods for the isolation and separation of extracellular vesicles from protein and lipid particles in human serum. Scientific Reports, 2020. 10(1): p. 1039.

13. Van Deun, J., et al., The impact of disparate isolation methods for extracellular vesicles on downstream RNA profiling. J Extracell Vesicles, 2014. 3.

14. Thery, C., et al., Minimal information for studies of extracellular vesicles 2018 (MISEV2018): a position statement of the International Society for Extracellular Vesicles and update of the MISEV2014 guidelines. J Extracell Vesicles, 2018. 7(1): p. 1535750.

15. Tian, Y., et al., Quality and efficiency assessment of six extracellular vesicle isolation methods by nano-flow cytometry. J Extracell Vesicles, 2020. 9(1): p. 1697028.

16. Veerman, R.E., et al., Molecular evaluation of five different isolation methods for extracellular vesicles reveals different clinical applicability and subcellular origin. Journal of Extracellular Vesicles, 2021. 10(9): p. e12128.

17. de Menezes-Neto, A., et al., Size-exclusion chromatography as a stand-alone methodology identifies novel markers in mass spectrometry analyses of plasma-derived vesicles from healthy individuals. J Extracell Vesicles, 2015. 4: p. 27378.

18. Gidlöf, O., et al., Proteomic profiling of extracellular vesicles reveals additional diagnostic biomarkers for myocardial infarction compared to plasma alone. Sci Rep, 2019. 9(1): p. 8991.

19. Askeland, A., et al., Mass-Spectrometry Based Proteome Comparison of Extracellular Vesicle Isolation Methods: Comparison of ME-kit, Size-Exclusion Chromatography, and High-Speed Centrifugation. Biomedicines, 2020. 8(8): p. 246.

20. Palviainen, M., et al., Extracellular vesicles from human plasma and serum are carriers of extravesicular cargo-Implications for biomarker discovery. PLoS One, 2020. 15(8): p. e0236439.

21. Cao, F., et al., Proteomics comparison of exosomes from serum and plasma between ultracentrifugation and polymer-based precipitation kit methods. Electrophoresis, 2019. 40(23-24): p. 3092–3098.

22. Jung, H.H., et al., Cytokine profiling in serum-derived exosomes isolated by different methods. Scientific Reports, 2020. 10(1): p. 14069.

23. Stranska, R., et al., Comparison of membrane affinity-based method with size-exclusion chromatography for isolation of exosome-like vesicles from human plasma. Journal of Translational Medicine, 2018. 16(1): p. 1.

24. Gutiérrez García, G., et al., Analysis of RNA yield in extracellular vesicles isolated by membrane affinity column and differential ultracentrifugation. PLoS One, 2020. 15(11): p. e0238545.

25. Peterka, O., et al., Lipidomic characterization of exosomes isolated from human plasma using various mass spectrometry techniques. Biochim Biophys Acta Mol Cell Biol Lipids, 2020. 1865(5): p. 158634.

26. Shtam, T., et al., Evaluation of immune and chemical precipitation methods for plasma exosome isolation. PLoS One, 2020. 15(11): p. e0242732.

27. Welton, J.L., et al., Ready-made chromatography columns for extracellular vesicle isolation from plasma. J Extracell Vesicles, 2015. 4: p. 27269.

28. Thompson, A.G., et al., CSF extracellular vesicle proteomics demonstrates altered protein homeostasis in amyotrophic lateral sclerosis. Clin Proteomics, 2020. 17: p. 31.

29. Subedi, P., et al., Comparison of methods to isolate proteins from extracellular vesicles for mass spectrometry-based proteomic analyses. Anal Biochem, 2019. 584: p. 113390.

30. Mi, H., et al., PANTHER version 14: more genomes, a new PANTHER GO-slim and improvements in enrichment analysis tools. Nucleic Acids Res, 2019. 47(D1): p. D419–D426.

31. Kim, D.K., et al., EVpedia: an integrated database of high-throughput data for systemic analyses of extracellular vesicles. J Extracell Vesicles, 2013. 2.

32. Kalra, H., et al., Vesiclepedia: a compendium for extracellular vesicles with continuous community annotation. PLoS Biol, 2012. 10(12): p. e1001450.

33. Simpson, R.J., H. Kalra, and S. Mathivanan, ExoCarta as a resource for exosomal research. J Extracell Vesicles, 2012. 1.

34. Jorgensen, M., et al., Extracellular Vesicle (EV) Array: microarray capturing of exosomes and other extracellular vesicles for multiplexed phenotyping. J Extracell Vesicles, 2013. 2.

35. Akawi, N., et al., Fat-Secreted Ceramides Regulate Vascular Redox State and Influence Outcomes in Patients With Cardiovascular Disease. Journal of the American College of Cardiology, 2021. 77(20): p. 2494–2513.

36. Serrano-Pertierra, E., et al., Characterization of Plasma-Derived Extracellular Vesicles Isolated by Different Methods: A Comparison Study. Bioengineering (Basel), 2019. 6(1).

37. Gámez-Valero, A., et al., Size-Exclusion Chromatography-based isolation minimally alters Extracellular Vesicles’ characteristics compared to precipitating agents. Sci Rep, 2016. 6: p. 33641.

38. Webber, J. and A. Clayton, How pure are your vesicles? J Extracell Vesicles, 2013. 2.

39. Théry, C., et al., Minimal information for studies of extracellular vesicles 2018 (MISEV2018): a position statement of the International Society for Extracellular Vesicles and update of the MISEV2014 guidelines. J Extracell Vesicles, 2018. 7(1): p. 1535750.

40. Skotland, T., et al., An emerging focus on lipids in extracellular vesicles. Adv Drug Deliv Rev, 2020. 159: p. 308–321.

41. Karimi, N., et al., Detailed analysis of the plasma extracellular vesicle proteome after separation from lipoproteins. Cell Mol Life Sci, 2018. 75(15): p. 2873–2886.

42. Yuana, Y., et al., Co-isolation of extracellular vesicles and high-density lipoproteins using density gradient ultracentrifugation. J Extracell Vesicles, 2014. 3.

43. Gidlof, O., et al., Proteomic profiling of extracellular vesicles reveals additional diagnostic biomarkers for myocardial infarction compared to plasma alone. Sci Rep, 2019. 9(1): p. 8991.

44. Bæk, R. and M.M. Jørgensen, Multiplexed Phenotyping of Small Extracellular Vesicles Using Protein Microarray (EV Array). Methods Mol Biol, 2017. 1545: p. 117–127.

45. Tóth, E., et al., Formation of a protein corona on the surface of extracellular vesicles in blood plasma. J Extracell Vesicles, 2021. 10(11): p. e12140.

46. Tian, B.M., et al., Human platelet lysate supports the formation of robust human periodontal ligament cell sheets. J Tissue Eng Regen Med, 2018. 12(4): p. 961–972.

47. Mathieu, M., et al., Specificities of exosome versus small ectosome secretion revealed by live intracellular tracking of CD63 and CD9. Nat Commun, 2021. 12(1): p. 4389.

48. Martínez-Beamonte, R., et al., Sphingomyelin in high-density lipoproteins: structural role and biological function. Int J Mol Sci, 2013. 14(4): p. 7716–41.

49. Sódar, B.W., et al., Low-density lipoprotein mimics blood plasma-derived exosomes and microvesicles during isolation and detection. Sci Rep, 2016. 6: p. 24316.

50. Izzo, C., F. Grillo, and E. Murador, Improved method for determination of high-density-lipoprotein cholesterol I. Isolation of high-density lipoproteins by use of polyethylene glycol 6000. Clin Chem, 1981. 27(3): p. 371–4.

51. Simonsen, J.B., What Are We Looking At? Extracellular Vesicles, Lipoproteins, or Both? Circ Res, 2017. 121(8): p. 920–922.

