## Supplemental Files for "Comparative and Integrated Analysis of Plasma Extracellular Vesicles Isolations Methods in Healthy Volunteers and Patients Following Myocardial Infarction"

Daan Paget et al.

### Supplemental Figure 1

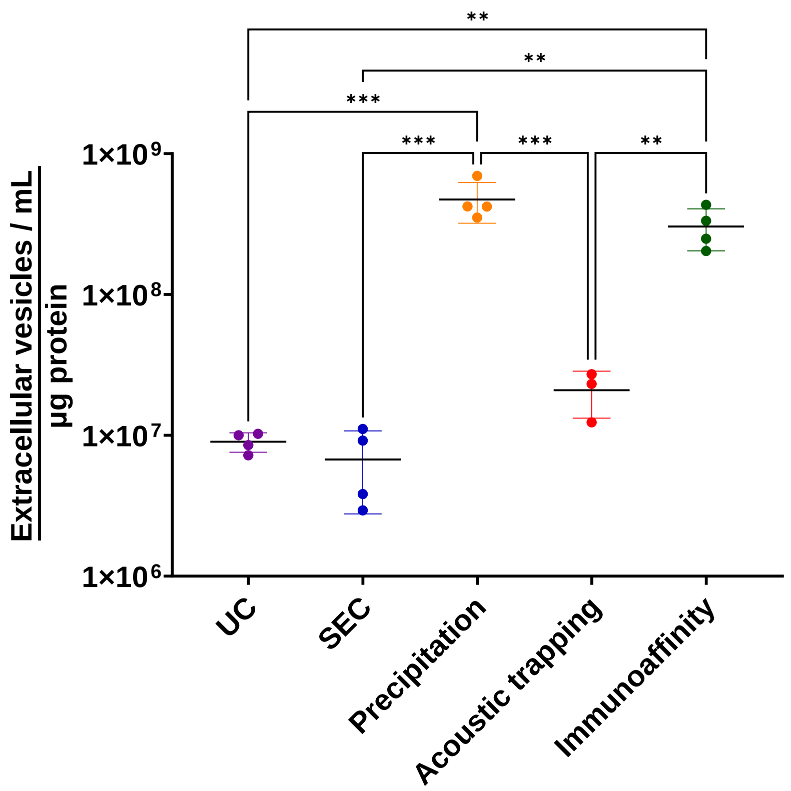

### Supplemental Figure 2

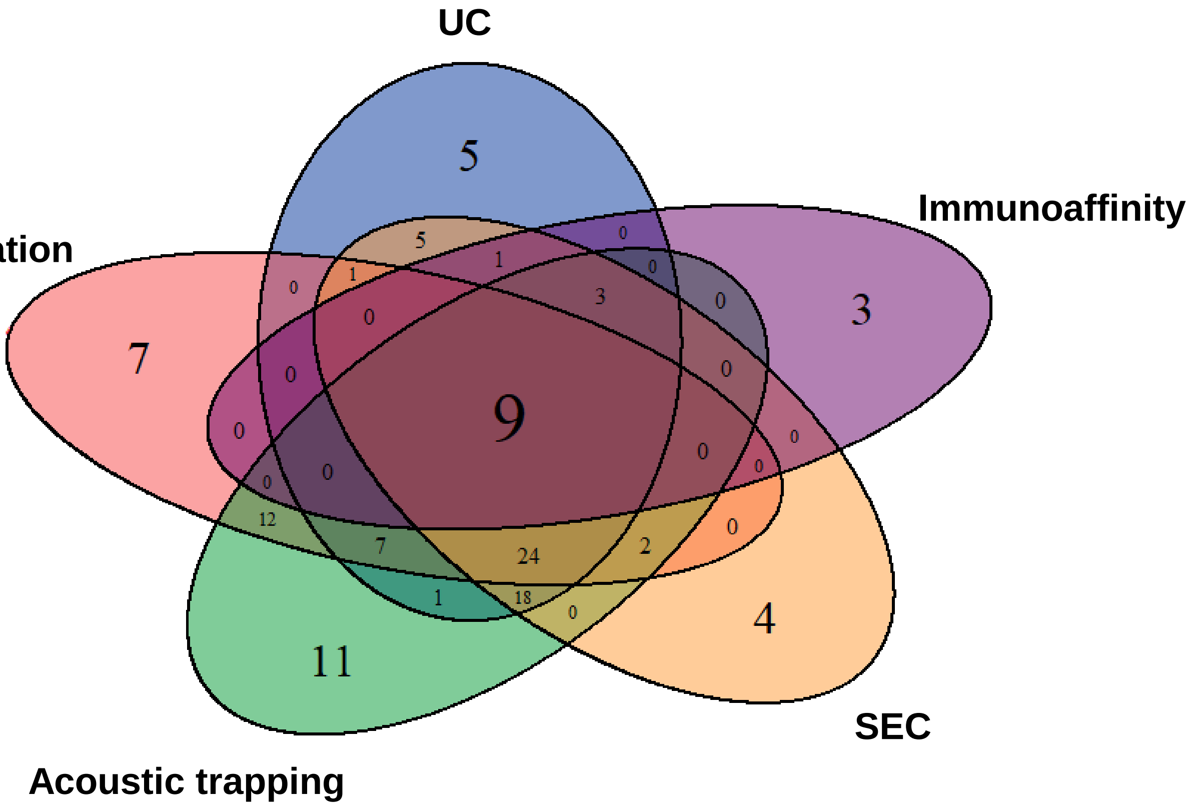

### Supplemental Figure 3

## A

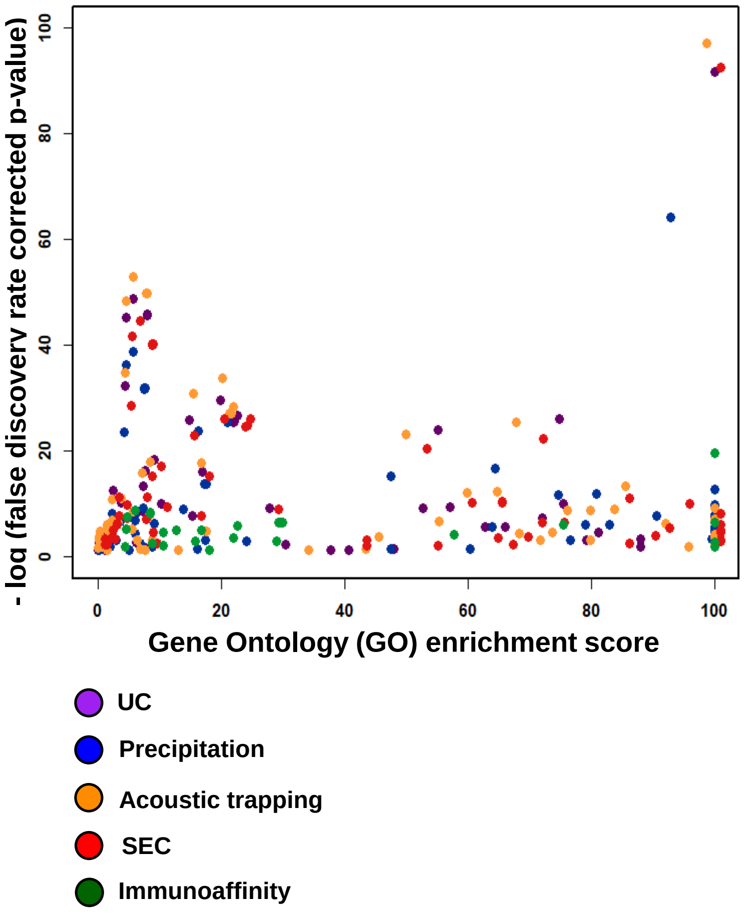


| UC - GO term | GO Enrichment Score | False discovery rate |
|---|---|---|
| 1. Blood microparticle | >100 | 1.46E-92 |
| 2. Extracellular space | 5.77 | 1.29E-49 |
| 3. Extracellular membrane-bounded organelle | 7.97 | 1.11E-46 |
| 4. Extracellular exosome | 8.05 | 1.17E-46 |
| 5. Extracellular organelle | 7.97 | 1.33E-46 |

| Precipitation - GO term | GO Enrichment Score | False discovery rate |
|---|---|---|
| 1. Blood microparticle | 92.92 | 4.79E-65 |
| 2. Extracellular space | 5.79 | 1.86E-39 |
| 3. Extracellular region | 4.64 | 3.87E-37 |
| 4. Extracellular membrane-bounded organelle | 7.52 | 1.19E-32 |
| 5. Extracellular exosome | 7.6 | 1.29E-32 |

| Acoustic trapping - GO term | GO Enrichment Score | False discovery rate |
|---|---|---|
| 1. Blood microparticle | 98.81 | 5.76E-98 |
| 2. Extracellular space | 5.73 | 9.74E-54 |
| 3. Extracellular exosome | 7.99 | 1.17E-50 |
| 4. Extracellular membrane-bounded organelle | 7.91 | 1.18E-50 |
| 5. Extracellular organelle | 7.91 | 1.41E-60 |

| SEC - GO term | GO Enrichment Score | False discovery rate |
|---|---|---|
| 1. Blood microparticle | >100 | 9.45E-93 |
| 2. Extracellular space | 5.82 | 9.30E-45 |
| 3. Extracellular region | 4.64 | 9.04E-42 |
| 4. Extracellular exosome | 7.97 | 2.61E-40 |
| 5. Extracellular membrane-bounded organelle | 7.88 | 2.62E-40 |

| Immunoaffinity - GO term. | GO Enrichment Score | False discovery rate |
|---|---|---|
| 1. Blood microparticle | >100 | 1.88E-20 |
| 2. Extracellular space | 6.08 | 1.77E-9 |
| 3. Extracellular membrane-bounded organelle | 8.42 | 4.30E-9 |
| 4. Extracellular organelle | 8.42 | 5.16E-9 |
| 5. Extracellular vesicle | 8.43 | 6.38E-9 |

## B

| | EVpedia | | Exocarta | | Vesiclepedia | |
|---|---|---|---|---|---|---|
| | % in common | P-value | % in common | P-value | % in common | P-value |
| Ultracentrifugation | 16.2 | 5.6E-12 | 6.8 | 7.8E-04 | 12.2 | 3.9E-08 |
| Precipitation | 9.7 | 3.1E-05 | 4.8 | 2.3E-02 | 4.8 | 2.3E-02 |
| Acoustic trapping | 10.3 | 1.6E-07 | 5.7 | 1.6E-03 | 6.9 | 2.1E-04 |
| SEC | 16.4 | 3.9E-11 | 9.0 | 4.8E-05 | 10.4 | 3.9E-06 |
| Immunoaffinity | 31.3 | 3.4E-07 | 18.8 | 4.8E-04 | 25.0 | 1.5E-05 |

### Supplemental Figure 4

| | UC | Precipitation | Acoustic trapping | SEC | Immunoaffinity |
|---|---|---|---|---|---|
| De Menezes-Neto et al. 2015 – ExoSpin | 55 / 74 | 43 / 62 | 61 / 87 | 52 / 67 | 10 / 16 |
| De Menezes-Neto et al. 2015 – SEC | 21 / 74 | 18 / 62 | 21 / 87 | 24 / 67 | 10 / 16 |
| Veerman et al. 2021 – ExoQuick | 56 / 74 | 47 / 62 | 66 / 87 | 50 / 67 | 10 / 16 |
| Veerman et al. 2021 – Izon35 | 51 / 74 | 40 / 62 | 59 / 87 | 47 / 67 | 10 / 16 |
| Veerman et al. 2021 – Izon70 | 39 / 74 | 28 / 62 | 41 / 87 | 41 / 67 | 9 / 16 |
| Veerman et al. 2021 - Optiprep | 55 / 74 | 45 / 62 | 65 / 87 | 50 / 67 | 10 / 16 |
| Veerman et al. 2021 – Exo-Easy | 55 / 74 | 46 / 62 | 66 / 87 | 50 / 67 | 10 / 16 |

### Supplemental Figure 5

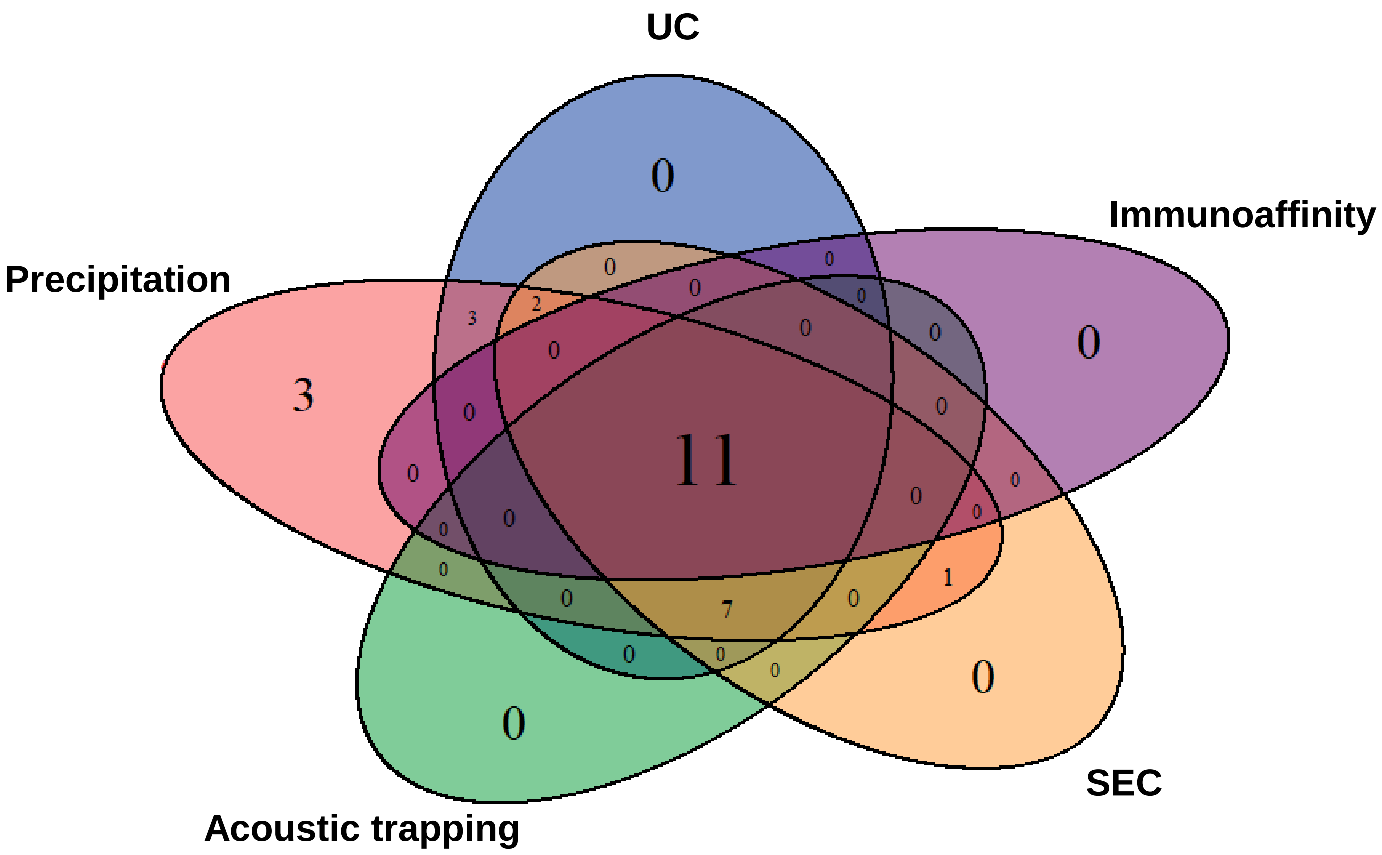

### Supplemental Figure 6

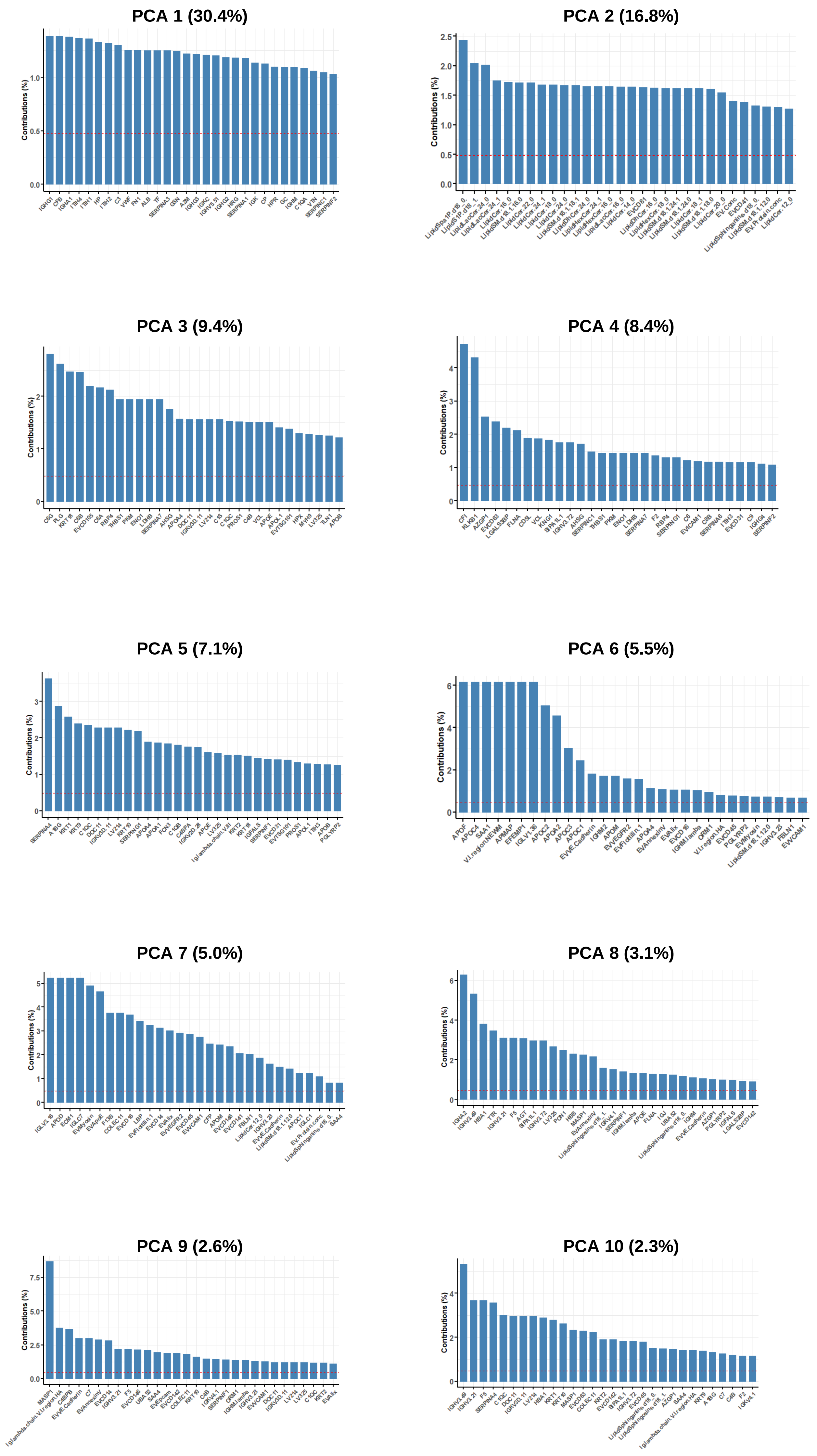

### Supplemental Figure 7

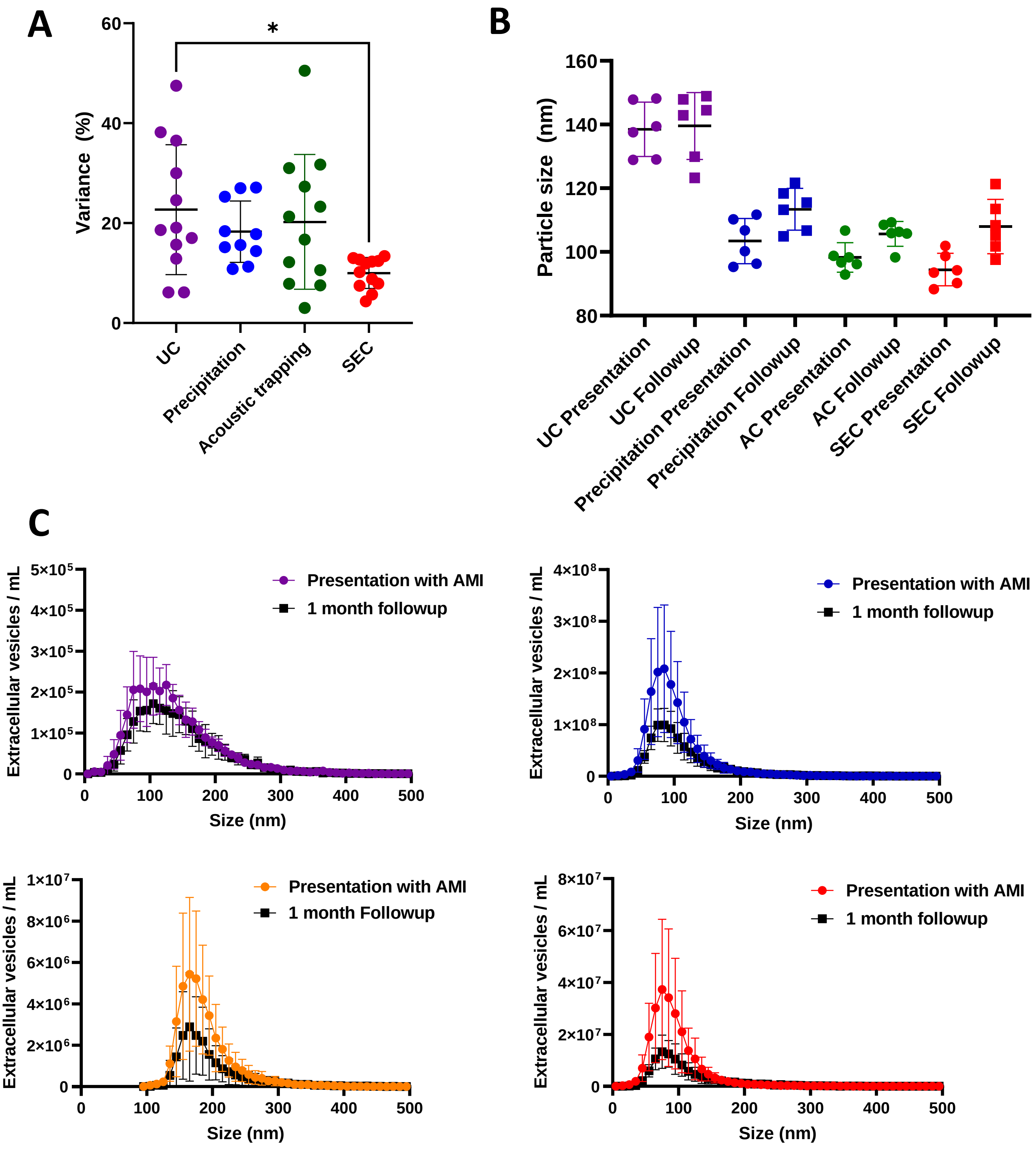

### Supplemental Figure 8

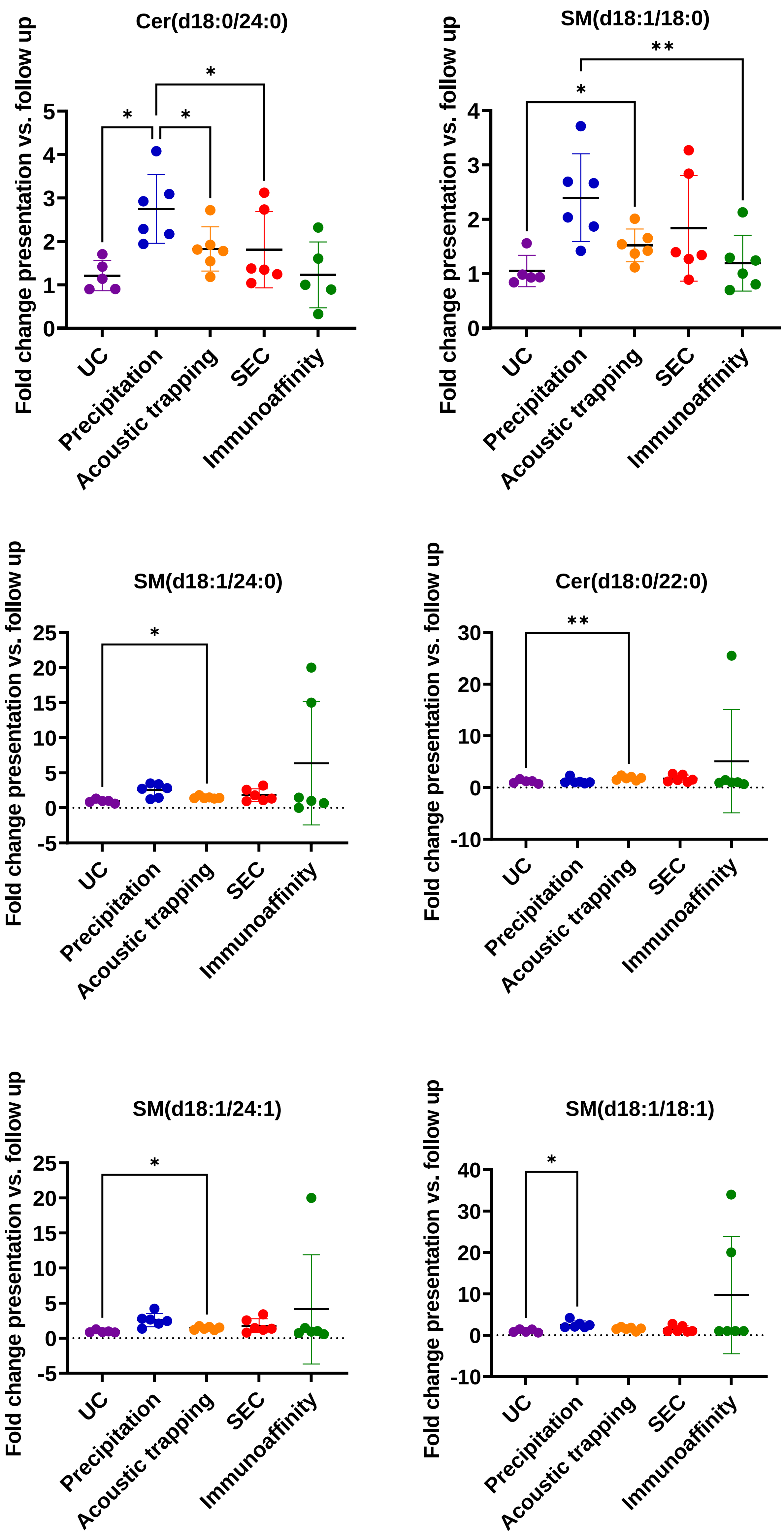

### Supplemental Figure 9

A

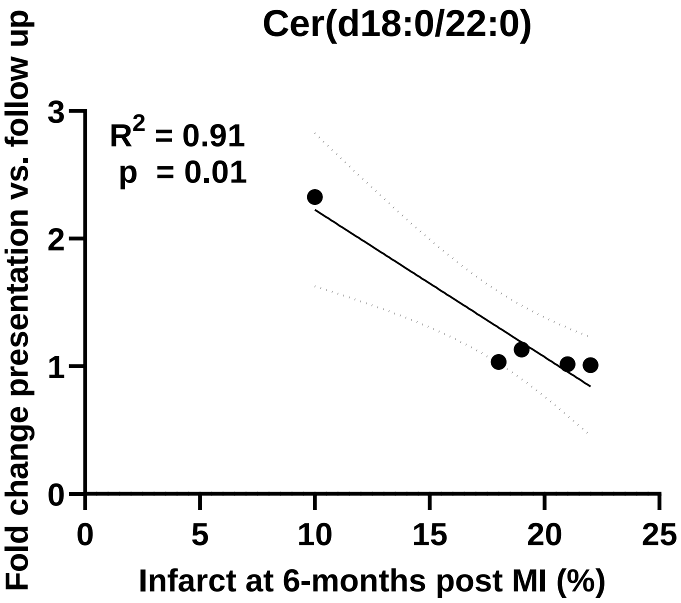


B

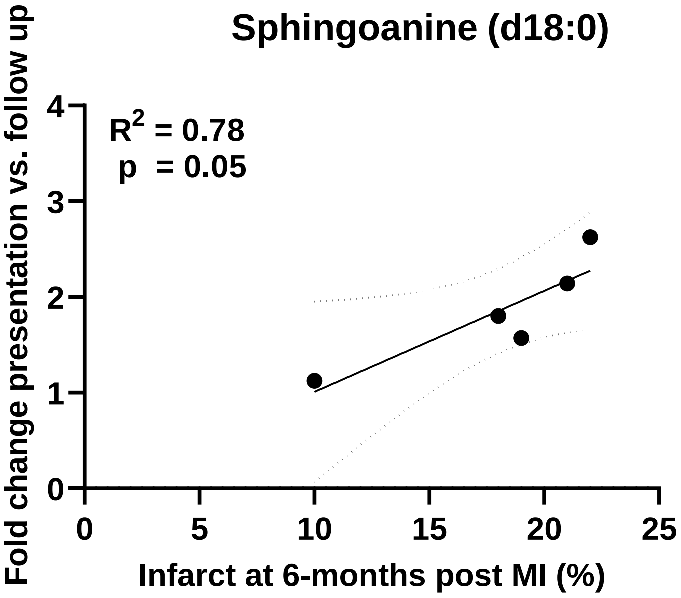


C

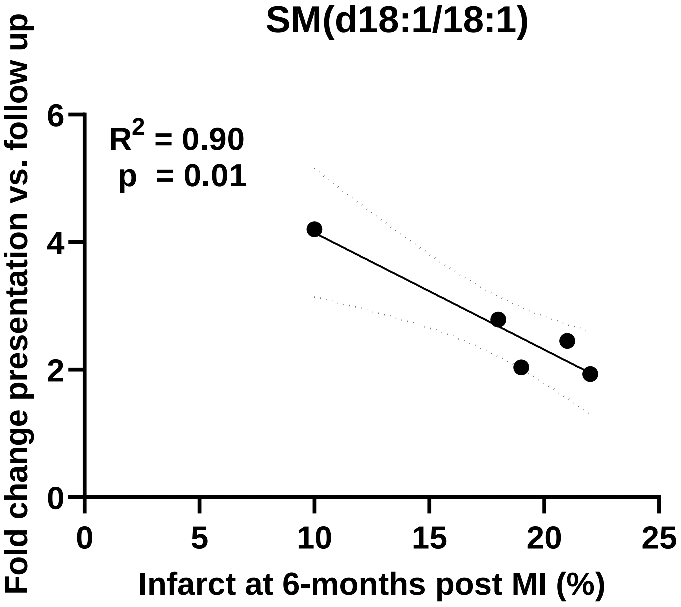
